# AI/ML-based prediction of TB treatment failure: A systematic review and meta-analysis

**DOI:** 10.64898/2026.04.16.26350453

**Authors:** Rogers Kamulegeya, Rose Nabatanzi, Derrick Semugenze, Faridah Mugala, Mercy Takuwa, Emmanuel Nasinghe, Denis Musinguzi, Sharon Namiiro, Andrew Katumba, Willy Ssengooba, Joyce Nakatumba-Nabende, Florence Nameere Kivunike, David Patrick Kateete

## Abstract

**Background:** Tuberculosis (TB) remains a leading cause of infectious disease mortality worldwide, and treatment failure contributes to ongoing transmission, drug resistance, and poor clinical outcomes. Artificial intelligence and machine learning approaches have attracted growing interest for predicting tuberculosis treatment outcomes, but the literature is heterogeneous and lacks a comprehensive synthesis.

**Methods:** We conducted a systematic review and meta-analysis of studies that developed or validated machine learning models to predict TB treatment failure. We searched PubMed/MEDLINE and Embase from January 2000 to October 2025. Studies were eligible if they developed, validated, or implemented an artificial intelligence or machine learning model for the prediction of TB treatment failure or a closely related poor outcome in patients receiving anti-TB treatment. Risk of bias was assessed using the Prediction model Risk Of Bias Assessment Tool. Random-effects meta-analysis was performed to pool area under the curve values, with subgroup analyses and meta-regression to explore heterogeneity.

**Results:** Thirty-four studies were included in the systematic review, of which 19 reported area under the curve values suitable for meta-analysis (total participants, 100,790). Studies were published between 2014 and 2025, with 91% published from 2019 onward. Tree-based methods were the most common algorithm family (52.9%), and multimodal models integrating three or more data types were used in 41.2% of studies. The pooled area under the curve was 0.836 (95% confidence interval 0.799–0.868), with substantial heterogeneity (I² = 97.9%). In subgroup analyses, studies including HIV-positive participants showed lower discrimination (pooled area under the curve 0.748) compared to those excluding them (0.924). Only eight studies (23.5%) performed external validation, and only one study (2.9%) was rated as low risk of bias overall, primarily due to methodological concerns in the analysis domain. Egger’s test suggested publication bias (p = 0.024). Major evidence gaps included underrepresentation of high-burden countries, HIV-affected populations, social determinants, pediatric TB, and extrapulmonary disease.

**Conclusions:** Machine learning models for predicting TB treatment failure show promising discrimination but are not yet ready for routine clinical implementation. Performance varies substantially across populations and settings, and methodological limitations, including inadequate validation, poor calibration assessment, and high risk of bias, limit confidence in current estimates. Future research should prioritize rigorous external validation, calibration assessment, and development in underrepresented populations, particularly HIV-affected and high-burden settings.

**Author Summary:** TB kills over a million people annually. While curable, treatment failure remains common and drives ongoing transmission and drug resistance. Researchers increasingly use artificial intelligence and machine learning to predict which patients will fail treatment, but it is unclear if these models are ready for clinical use. We reviewed 34 studies including nearly 1.1 million participants from 22 countries. On average, models correctly distinguished patients who would fail treatment from those who would not 84% of the time, a performance generally considered good. However, this average hid enormous variation. Models developed in populations including HIV-positive people performed substantially worse, suggesting prediction is harder with HIV co-infection. Worryingly, only one study used high-quality methods; 97% had serious flaws in handling missing data, checking calibration, or testing in new populations. Only eight studies validated their models in different settings. To conclude, we found that machine learning is promising in predicting TB treatment failure, but it is not ready for clinical use. Researchers should prioritize validation in high-burden settings, include social determinants, and improve methodological rigor before these tools can help patients.

## Introduction

Tuberculosis, caused by *Mycobacterium tuberculosis*, remains a major global health challenge and one of the leading causes of death from a single infectious agent worldwide [1]. In 2023, an estimated 10.8 million people developed tuberculosis and 1.25 million died from the disease [1]. Although tuberculosis is curable with standard antibiotic regimens, treatment failure remains clinically and programmatically important because it is associated with persistent infectiousness, prolonged morbidity, relapse, amplification of drug resistance, and continued community transmission [2–7]. These risks are particularly consequential in settings with constrained health systems, delayed diagnosis, high levels of drug-resistant tuberculosis, or substantial HIV coinfection.

The determinants of unsuccessful tuberculosis treatment outcomes are multifactorial and interconnected. Pathogen-related drivers include drug resistance profile and bacillary burden [6,7]. Host-related drivers include immune status, HIV coinfection, diabetes, malnutrition, and other comorbidities [8,9]. Health system factors, such as continuity of drug supply, treatment access, and quality of monitoring, also shape outcomes, while social and behavioral determinants including poverty, overcrowding, stigma, food insecurity, substance use, and treatment adherence further influence risk [5,10–12]. These determinants interact in complex and often nonlinear ways, making prognosis difficult to capture using simple rule-based or narrowly specified analytic approaches [13,14].

Traditional prognostic approaches in tuberculosis have relied mainly on standard statistical models built from a limited number of clinical and laboratory predictors. These models remain valuable and interpretable, but they may be less able to capture nonlinear relationships, high-dimensional interactions, and heterogeneity across diverse data sources [13–17]. Artificial intelligence and machine learning approaches have therefore attracted growing interest as tools for prediction in tuberculosis care. In principle, such models can integrate demographic, clinical, microbiological, radiological, pharmacokinetic, genomic, and socioeconomic information to generate individualized risk estimates [9–17].

Over the past decade, a growing body of work has applied artificial intelligence and machine learning approaches to predict treatment failure and other poor tuberculosis treatment outcomes [18–21]. However, this literature remains heterogeneous and fragmented. Studies differ in the populations included, ranging from drug-susceptible to multidrug-resistant and extensively drug-resistant tuberculosis; in epidemiologic context, including high tuberculosis burden, tuberculosis and HIV burdened, and lower-burden settings; in predictor domains; in model families; in validation strategies; and in the outcomes and performance metrics reported [20–23]. As a result, it remains unclear how well these models perform overall, which data modalities are most informative, and whether the published evidence is sufficiently robust to support clinical translation.

These questions are especially important in settings where tuberculosis burden is highest. A model developed in a lower-burden or data-rich environment may not transport well to high-burden settings, where health system constraints, comorbidity patterns, treatment pathways, and baseline risk differ substantially [24,25]. Similarly, populations in tuberculosis and HIV burdened settings may have distinct risk structures that are not captured by models trained in predominantly HIV-negative cohorts. It also remains uncertain whether the marginal predictive value of more expensive inputs, such as genomics or pharmacokinetic measurements, justifies their collection in low-resource environments. Furthermore, despite the established importance of social determinants in tuberculosis outcomes, these factors appear to be underrepresented in published prediction models.

To our knowledge, no previous systematic review has comprehensively synthesized artificial intelligence and machine learning models for tuberculosis treatment failure prediction while also quantitatively pooling discrimination, examining heterogeneity across geographic and epidemiologic contexts, and appraising methodological quality using the Prediction model Risk Of Bias Assessment Tool [22,23]. We therefore conducted a systematic review and meta-analysis to evaluate the predictive performance of artificial intelligence and machine learning models for tuberculosis treatment failure; to compare performance across high versus lower tuberculosis burden settings, tuberculosis and HIV burdened settings, and income contexts; to characterize commonly used predictor domains and data modalities; to assess methodological quality and risk of bias; to evaluate validation rigor and generalizability; and to identify knowledge gaps to inform future model development and implementation.

## Results

### Study selection

The systematic search of PubMed/MEDLINE and Embase identified 1,672 records. After removal of 1,344 records during initial filtering based on study focus, language, publication type, and other exclusion criteria, 328 records remained. Following the removal of 11 duplicates, 317 abstracts were screened independently by three reviewers. Of these, 265 were excluded because they did not address prediction modeling for tuberculosis treatment outcomes. Fifty-two full-text articles were assessed for eligibility, and 18 were excluded because they used only conventional statistical approaches without a machine learning component (n=15) or did not evaluate treatment failure as an outcome (n=3). Thirty-four studies met the inclusion criteria for the systematic review, of which 19 reported area under the receiver operating characteristic curve (AUC) values suitable for quantitative meta-analysis (S1 Figure – PRISMA Flow diagram).

AUC quantifies how well a model distinguishes between patients who experience treatment failure and those who do not, with values ranging from 0.5 (no discrimination beyond chance) to 1.0 (perfect discrimination).

### Study characteristics

#### Temporal and geographic distribution

The included studies were published between 2014 and 2025, with a marked increase in recent years. Only three studies appeared before 2019, whereas 31 studies (91.2%) were published from 2019 onward, reflecting rapidly growing interest in artificial intelligence and machine learning approaches for predicting tuberculosis treatment outcomes (Figure 1).

**Figure 1.**
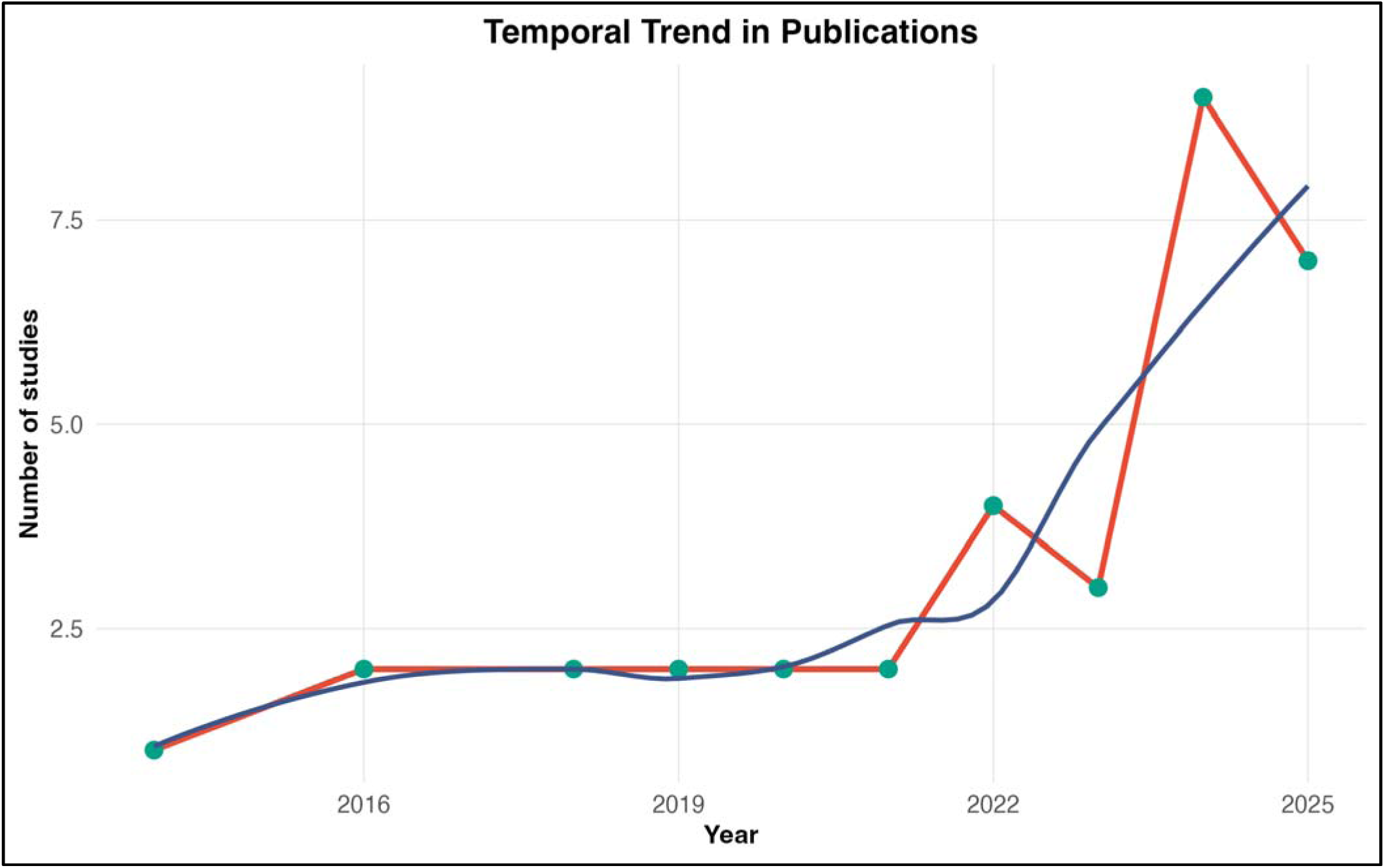
Temporal Trend in Publications. Bar chart showing the annual number of included studies published between 2014 and 2025. The number of publications remained low (1–2 per year) until 2020, then increased markedly, with 6 studies published in 2024 and 7 studies in 2025. This trend indicates a rapidly growing research interest in machine learning applications for tuberculosis treatment outcome prediction.

Geographically, studies were conducted across 22 countries spanning Asia, Africa, Europe, and the Americas (Table 1; Figure 2), but the distribution was uneven. China contributed the largest number of studies (n=10, 29.4%), followed by India (n=5, 14.7%), while multinational collaborations accounted for four studies (11.8%). Several countries contributed only one study.

**Figure 2.**
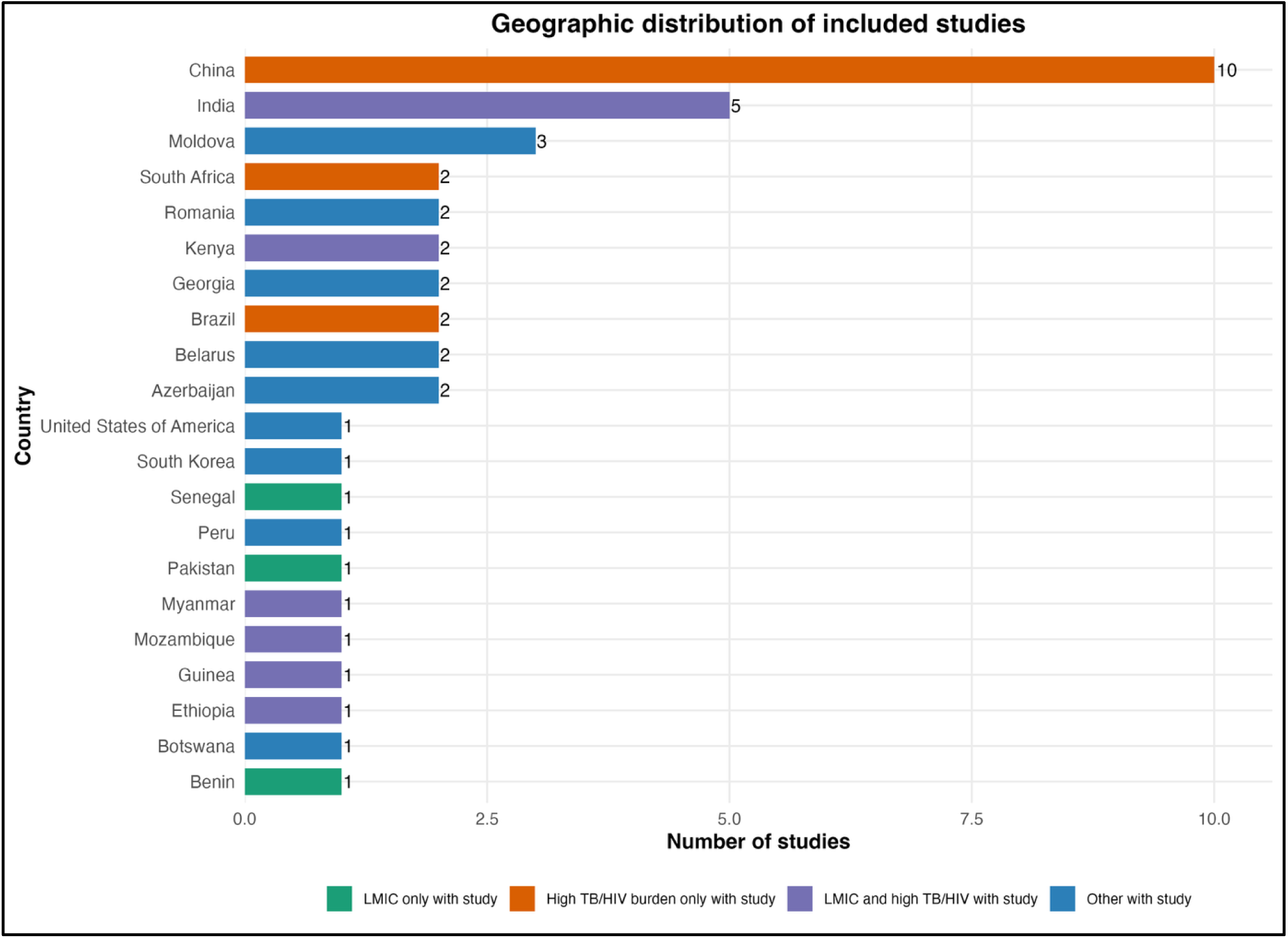
Geographic Distribution by Country Classification. Bar chart showing the distribution of included studies across countries, categorized by low- and middle-income country status and high tuberculosis/HIV burden status. China contributed the largest number of studies (n=10), followed by India (n=4). Many countries with high tuberculosis or tuberculosis/HIV burden contributed only one study each (e.g., Ethiopia, Kenya, Mozambique, South Africa), while several high-burden countries had no eligible studies. The “Other” category includes countries such as the United States, South Korea, and others not classified as low- and middle-income or high tuberculosis/HIV burden.

**Table 1.**
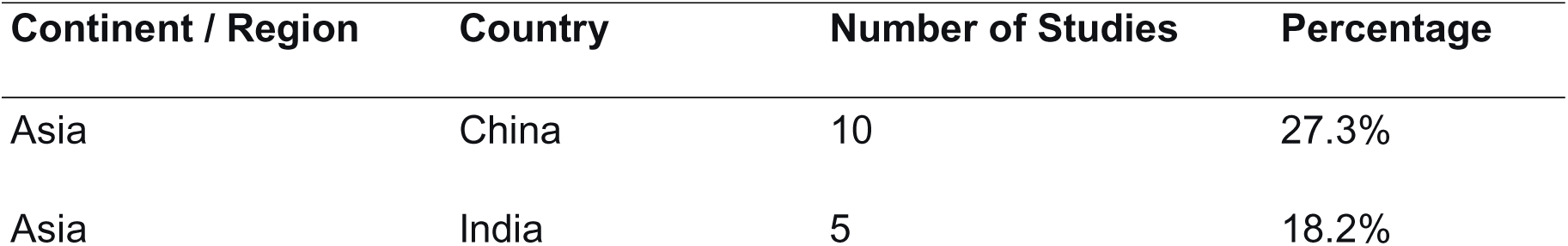
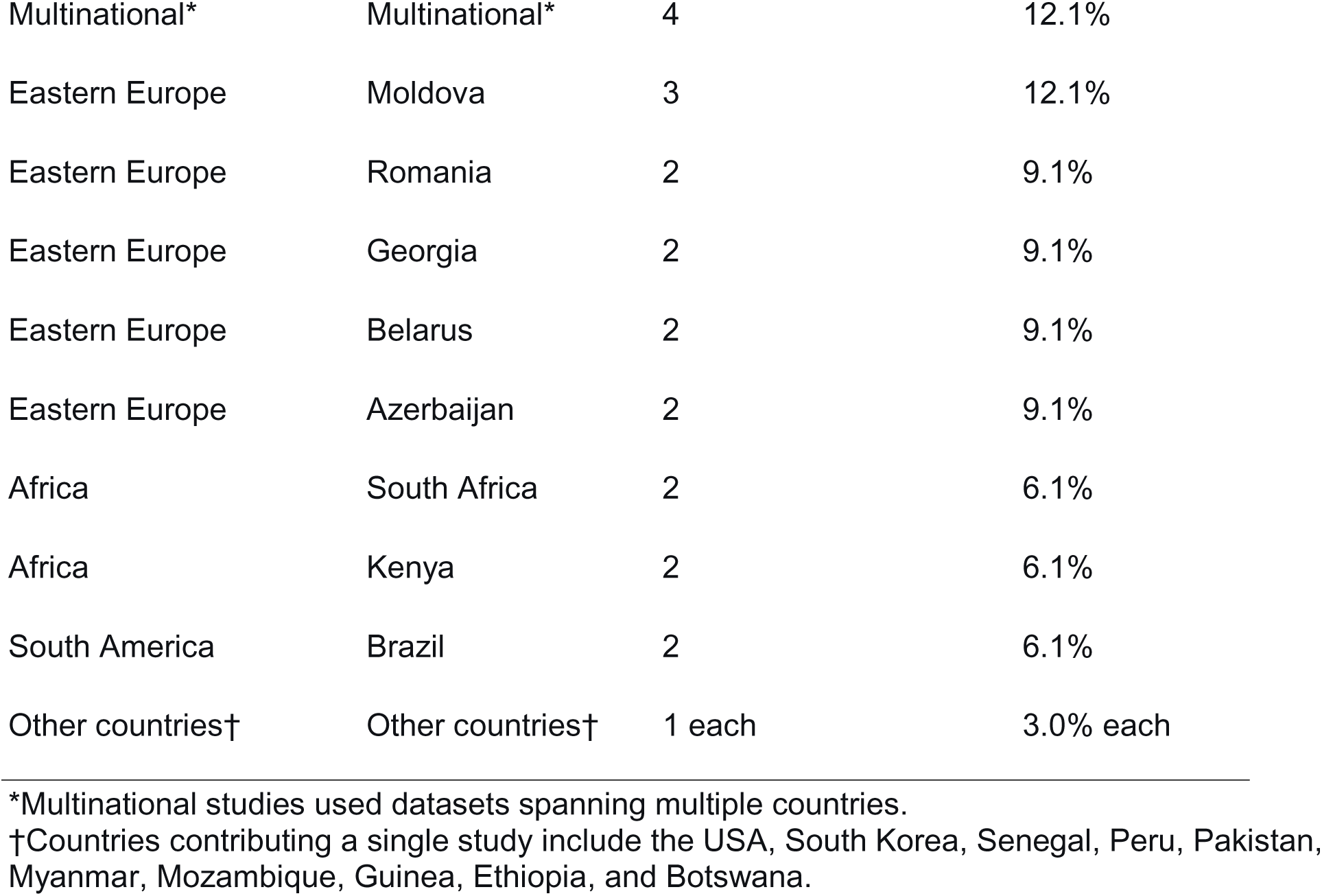
Geographic Distribution of Included Studies.

| Continent / Region | Country | Number of Studies | Percentage |
| --- | --- | --- | --- |
| Asia | China | 10 | 27.3% |
| Asia | India | 5 | 18.2% |
| Multinational* | Multinational* | 4 | 12.1% |
| Eastern Europe | Moldova | 3 | 12.1% |
| Eastern Europe | Romania | 2 | 9.1% |
| Eastern Europe | Georgia | 2 | 9.1% |
| Eastern Europe | Belarus | 2 | 9.1% |
| Eastern Europe | Azerbaijan | 2 | 9.1% |
| Africa | South Africa | 2 | 6.1% |
| Africa | Kenya | 2 | 6.1% |
| South America | Brazil | 2 | 6.1% |
| Other countries† | Other countries† | 1 each | 3.0% each |

\*Multinational studies used datasets spanning multiple countries.
†Countries contributing a single study include the USA, South Korea, Senegal, Peru, Pakistan, Myanmar, Mozambique, Guinea, Ethiopia, and Botswana.

Mapping study locations against World Bank income classification and WHO high TB/HIV burden countries revealed substantial gaps (Figure 3). Although many studies originated from LMICs, relatively few were conducted in countries simultaneously classified as LMIC and high TB/HIV burden settings, and many high-burden countries had no eligible studies. This uneven geographic distribution may partly explain the substantial heterogeneity observed in model performance across studies.

**Figure 3.**
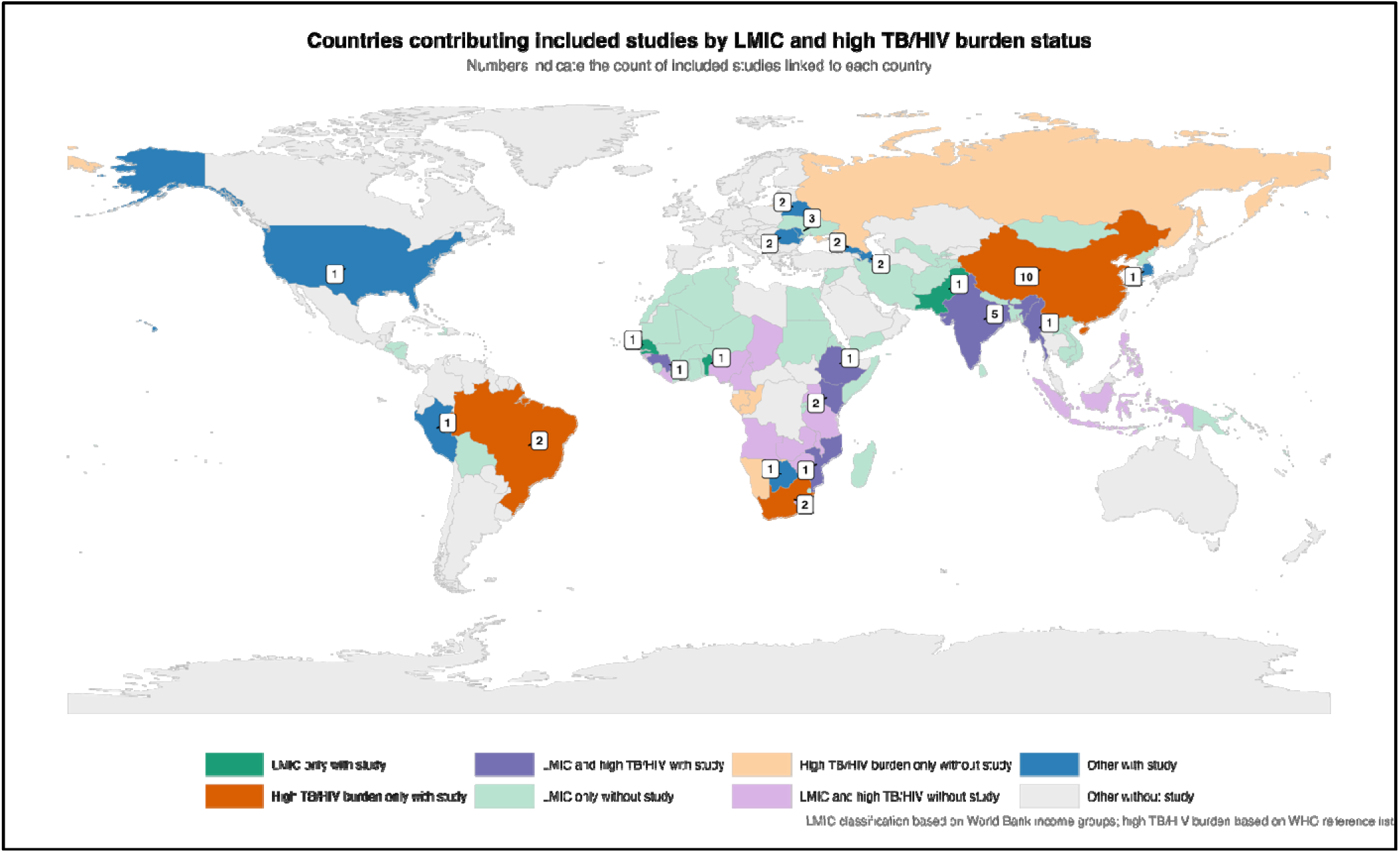
World Map of Study Distribution. World map showing the geographic distribution of included studies, with numbers indicating the count of studies linked to each country. The map highlights the concentration of studies in China and India, with sparse representation from sub-Saharan Africa, Southeast Asia, and other high-burden regions. Countries with no eligible studies are shown in light gray.

#### Study design, setting, and sample size

Most studies used retrospective observational designs (24/34, 70.6%), while only three studies used prospective cohorts (Table 2). Sample sizes varied widely, ranging from 28 to 665,883 participants (median 551.5, IQR 197–4,139). The included studies were conducted in diverse settings, including tertiary referral hospitals, district and community facilities, national tuberculosis program datasets, and multicenter or international databases. This diversity broadens the clinical scope of the review, but it also contributes to heterogeneity in patient mix, predictor availability, and outcome ascertainment.

**Table 2.** Summary of study design, setting, and sample size characteristics of the included studies.

| Characteristic | Category | Studies, n (%) |
| --- | --- | --- |
| Study design | Retrospective cohort / retrospective observational | 24 (70.6) |
|  | Prospective cohort / prospective clinical study | 3 (8.8) |
|  | Nested case–control or case–control | 3 (8.8) |
|  | Mathematical / transmission modeling studies | 2 (5.9) |
|  | Longitudinal pilot / validation study | 1 (2.9) |
|  | Other hybrid designs | 1 (2.9) |
| Study setting | Single-country datasets | 30 (88.2) |
|  | Multinational datasets | 4 (11.8) |
| Sample size | Median sample size (IQR) | 551.5 (197–4,139) |
|  | Minimum sample size | 28 |
|  | Maximum sample size | 665,883 |

#### Population characteristics

Population descriptors varied across studies (Table 3; Figure 4). HIV status was reported in 20 of 34 studies (58.8%), with 10 including HIV-positive participants and 4 explicitly excluding them, while 14 studies did not report HIV status. Drug resistance profiles were also heterogeneous, with studies focusing on drug-susceptible, drug-resistant, or mixed populations. Most studies investigated pulmonary tuberculosis, while a smaller proportion included both pulmonary and extrapulmonary disease or focused exclusively on extrapulmonary cases. Diabetes was the most reported non-HIV comorbidity. These differences in study populations likely contributed to the heterogeneity observed in model performance across studies.

**Figure 4.**
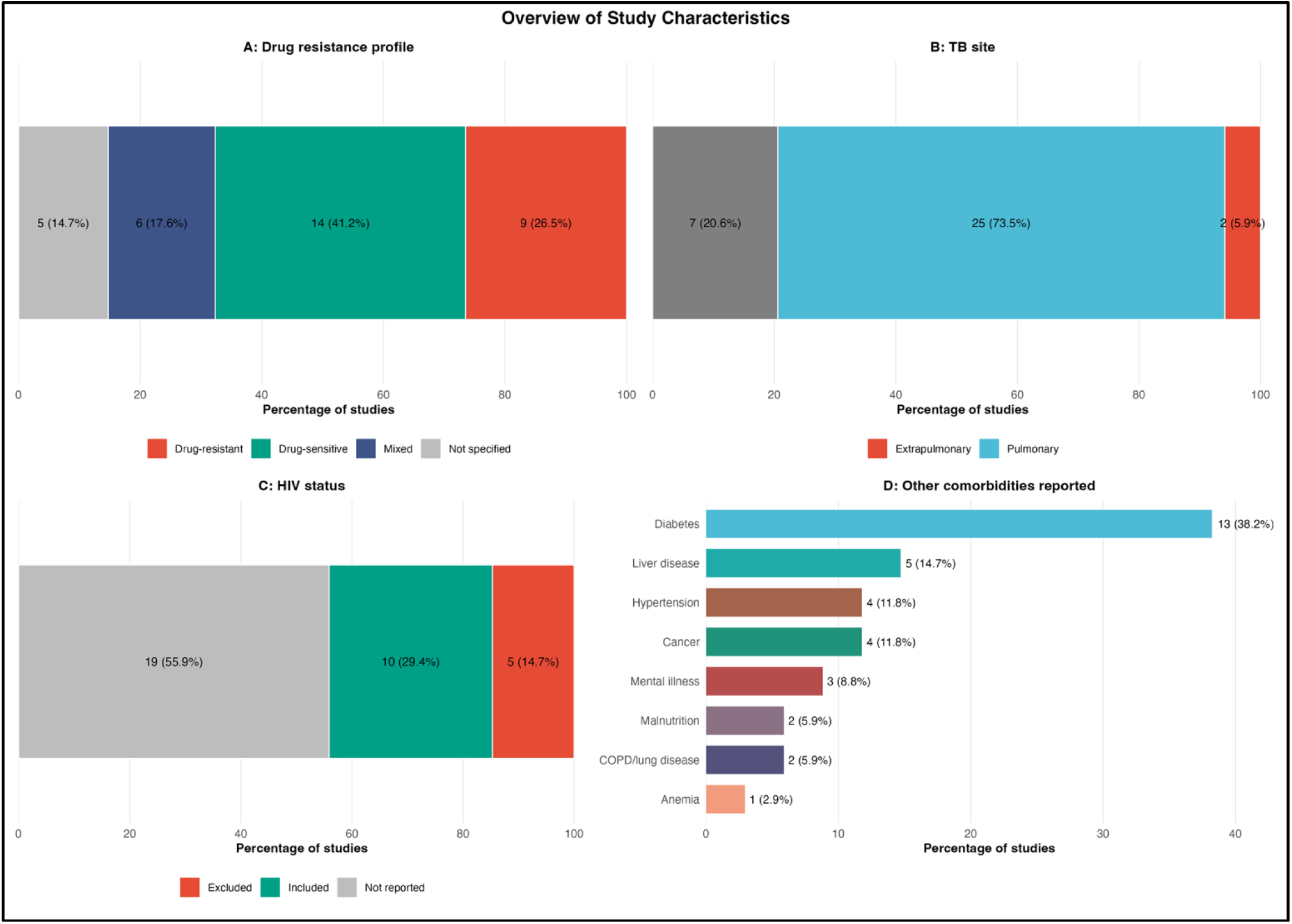
Overview of Study Population Characteristics. Four-panel figure summarizing key population characteristics of the 34 included studies. **Panel A: Drug Resistance Profile.** Fourteen studies (41.2%) focused on drug-susceptible tuberculosis, nine (26.5%) on drug-resistant tuberculosis, nine (26.5%) on mixed populations, and two (5.9%) did not specify resistance profile. **Panel B: Tuberculosis Site.** Twenty-five studies (73.5%) focused exclusively on pulmonary tuberculosis, while nine (26.5%) included both pulmonary and extrapulmonary cases. No studies focused exclusively on extrapulmonary tuberculosis. **Panel C: HIV Status.** Ten studies (29.4%) included HIV-positive participants, four (11.8%) explicitly excluded HIV-positive individuals, and 14 (41.2%) did not report HIV status. **Panel D: Other Comorbidities Reported.** Diabetes was the most frequently reported non-HIV comorbidity (13 studies, 38.2%), followed by liver disease (5, 14.7%), hypertension (4, 11.8%), cancer (4, 11.8%), and mental illness (3, 8.8%).

**Table 3.** Population Characteristics Reported in Included Studies (N = 34)

| Population Descriptor | Category | Studies, n (%) |
| --- | --- | --- |

| **Population Descriptor** | **Category** | **Studies, n (%)** |
| --- | --- | --- |
| **HIV status reporting** | HIV-positive participants included | 10 (29.4) |
|  | HIV-positive participants excluded | 4 (11.8) |
|  | HIV status reported but inclusion not specified | 6 (17.6) |
|  | HIV status not reported | 14 (41.2) |
| **Drug resistance profile** | Drug-susceptible tuberculosis | 14 (41.2) |
|  | Drug-resistant tuberculosis | 9 (26.5) |
|  | Mixed resistance populations | 9 (26.5) |
|  | Not specified | 2 (5.9) |
| **Tuberculosis disease site** | Pulmonary tuberculosis only | 25 (73.5) |
|  | Pulmonary and extrapulmonary tuberculosis | 7 (20.6) |
|  | Exclusively extrapulmonary tuberculosis | 2 (5.9) |
| **Non-HIV comorbidities reported\*** | Diabetes mellitus | 13 (38.2) |
|  | Liver disease | 5 (14.7) |
|  | Hypertension | 4 (11.8) |
|  | Cancer | 4 (11.8) |
|  | Mental illness | 3 (8.8) |

#### Outcome definitions

Outcome definitions varied substantially across studies (Table 4). The most common definition was WHO-aligned bacteriological treatment failure, based on persistent smear or culture positivity at treatment completion or after five months of therapy. Other studies used composite unfavorable outcomes that combined treatment failure with events such as death or loss to follow-up, while several defined “failure” using lack of sputum culture conversion at predefined treatment milestones.

**Table 4.**
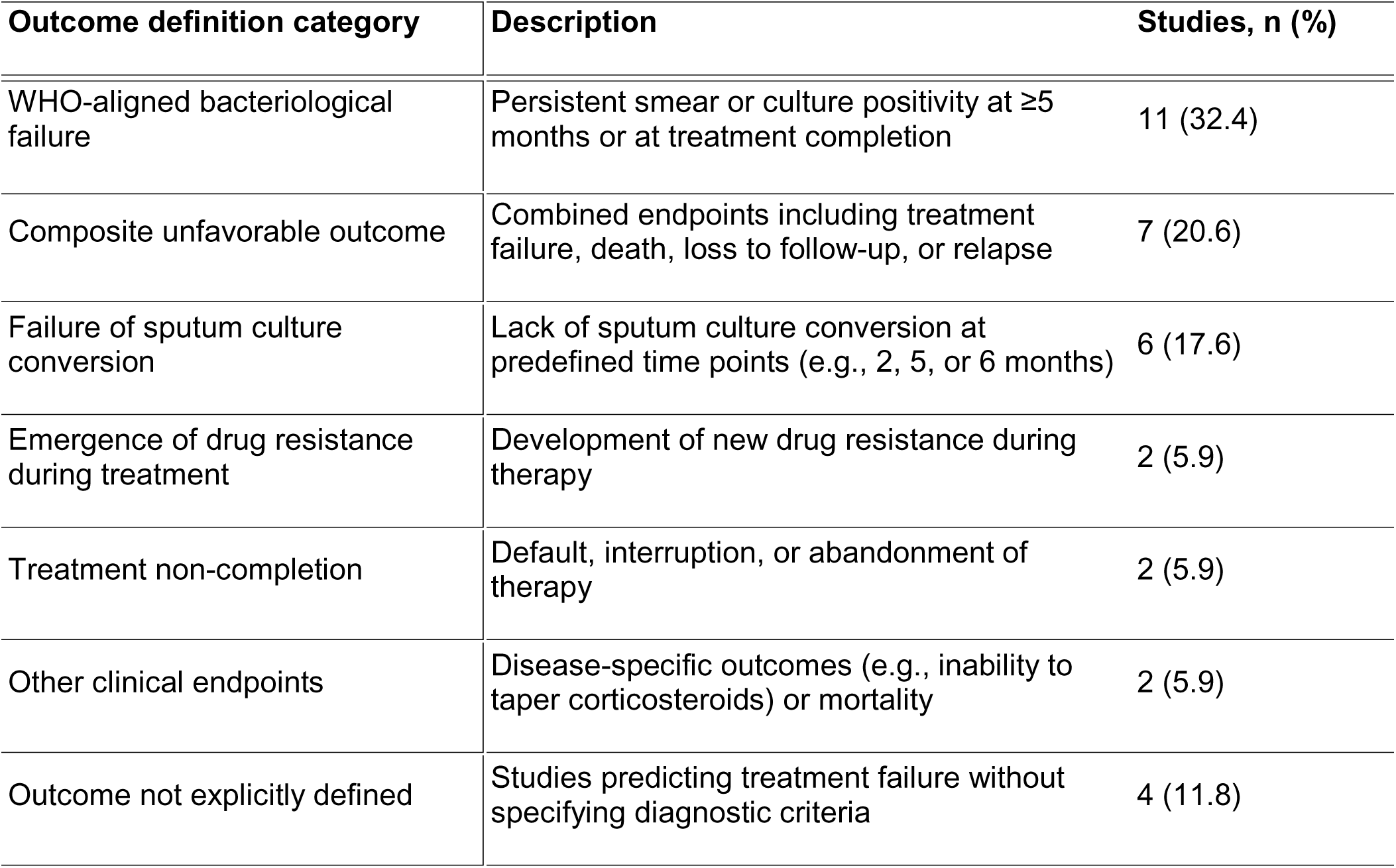
Operational definitions of treatment failure outcomes used in the included studies (N = 34)

Less frequently, studies defined treatment failure using outcomes such as development of drug resistance during therapy, treatment non-completion, disease-specific clinical endpoints, or all-cause mortality. In addition, several studies did not clearly specify the outcome definition used.

Overall, this variation indicates that models described as predicting “treatment failure” were often trained on different clinical endpoints, which limits comparability across studies and likely contributed to the high heterogeneity observed in the pooled analyses.

### Model characteristics

#### Machine learning algorithms and algorithm families

A wide range of machine learning algorithms were evaluated across the included studies. Among the 30 studies that reported a best-performing model, tree-based approaches were most frequently identified. Random forest was the most selected best-performing algorithm, appearing in 9 studies (30.0%). Decision trees or classification and regression tree (CART) models were reported as best in 5 studies (16.7%), while gradient boosting approaches such as XGBoost and LightGBM were best in 4 studies (13.3%). Logistic regression models, including regularized variants, were the best-performing models in 4 studies (13.3%). Neural networks were reported as best in 3 studies (10.0%), and support vector machines in 2 studies (6.7%). Other approaches, including k-nearest neighbors, elastic net models, and ensemble methods, accounted for the remaining 3 studies (10.0%).

When grouped into broader methodological families, tree-based methods, including random forest, decision trees, and gradient boosting, dominated the literature, appearing in 18 studies (52.9%). Regression-based models (logistic regression, LASSO, and elastic net) were used in 8 studies (23.5%), neural network or deep learning models in 4 studies (11.8%), and other machine learning approaches such as support vector machines and k-nearest neighbors in 4 studies (11.8%).

The predominance of tree-based methods likely reflects their practical advantages in clinical datasets, including the ability to accommodate missing data, capture nonlinear relationships, and model complex interactions among clinical, demographic, and microbiological variables. Consistent with this methodological preference, many of the highest-performing models reported in the included studies were based on tree-based algorithms, suggesting that these approaches may be particularly well suited to the heterogeneous and multimodal data commonly encountered in tuberculosis treatment outcome prediction.

#### Data modalities and multimodal modeling

Studies differed substantially in the types of data used to develop prediction models. Predictors were broadly grouped into several domains, including clinical and demographic characteristics (such as age, sex, symptoms, and medical history), laboratory biomarkers (blood tests and inflammatory markers), microbiological data (smear microscopy, culture, GeneXpert, and drug susceptibility testing), radiological or imaging features (e.g., chest X-ray or computed tomography), pharmacokinetic measures (such as drug concentrations and exposure metrics), genomic or other omics data, and social or behavioral factors including education, employment, income, alcohol use, smoking, and substance use.

Most studies incorporated information from multiple domains rather than relying on a single type of data. Figure 5 illustrates the combinations of predictor modalities used across studies, highlighting that clinical and demographic variables formed the backbone of many models and were frequently combined with laboratory or pharmacokinetic data. Other models incorporated imaging or omics features, although these approaches were less common.

**Figure 5.**
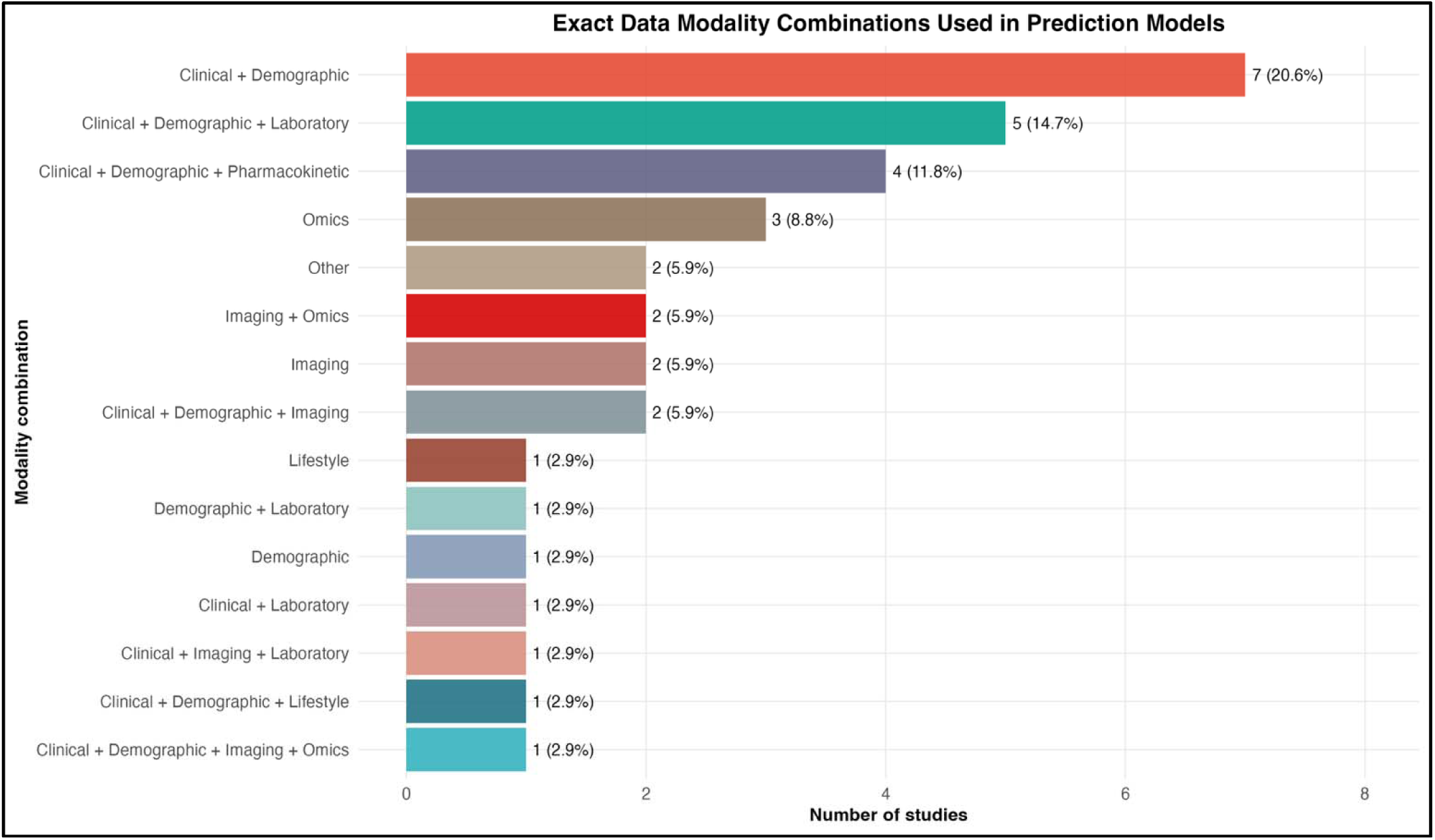
Data Modality Combinations Used in Prediction Models. Horizontal bar chart showing the specific combinations of data modalities used across the 34 included studies. The most frequent combinations were clinical plus demographic data only (7 studies, 20.6%), clinical plus demographic plus laboratory data (5, 14.7%), clinical plus demographic plus pharmacokinetic data (4, 11.8%), and omics data only (3, 8.8%). The variety of combinations reflects the diversity of approaches to multimodal modeling in tuberculosis treatment outcome prediction.

Model discrimination varied modestly according to the number of modalities used. Among the 19 studies reporting AUC values suitable for comparison, single-modality models achieved a mean AUC of 0.802, dual-modality models achieved 0.830, and multimodal models integrating three or more data sources achieved 0.841 (Figure 6). While the increase in discrimination was relatively small, the overall pattern suggests that integrating multiple data sources may provide incremental improvements in predictive performance.

**Figure 6.**
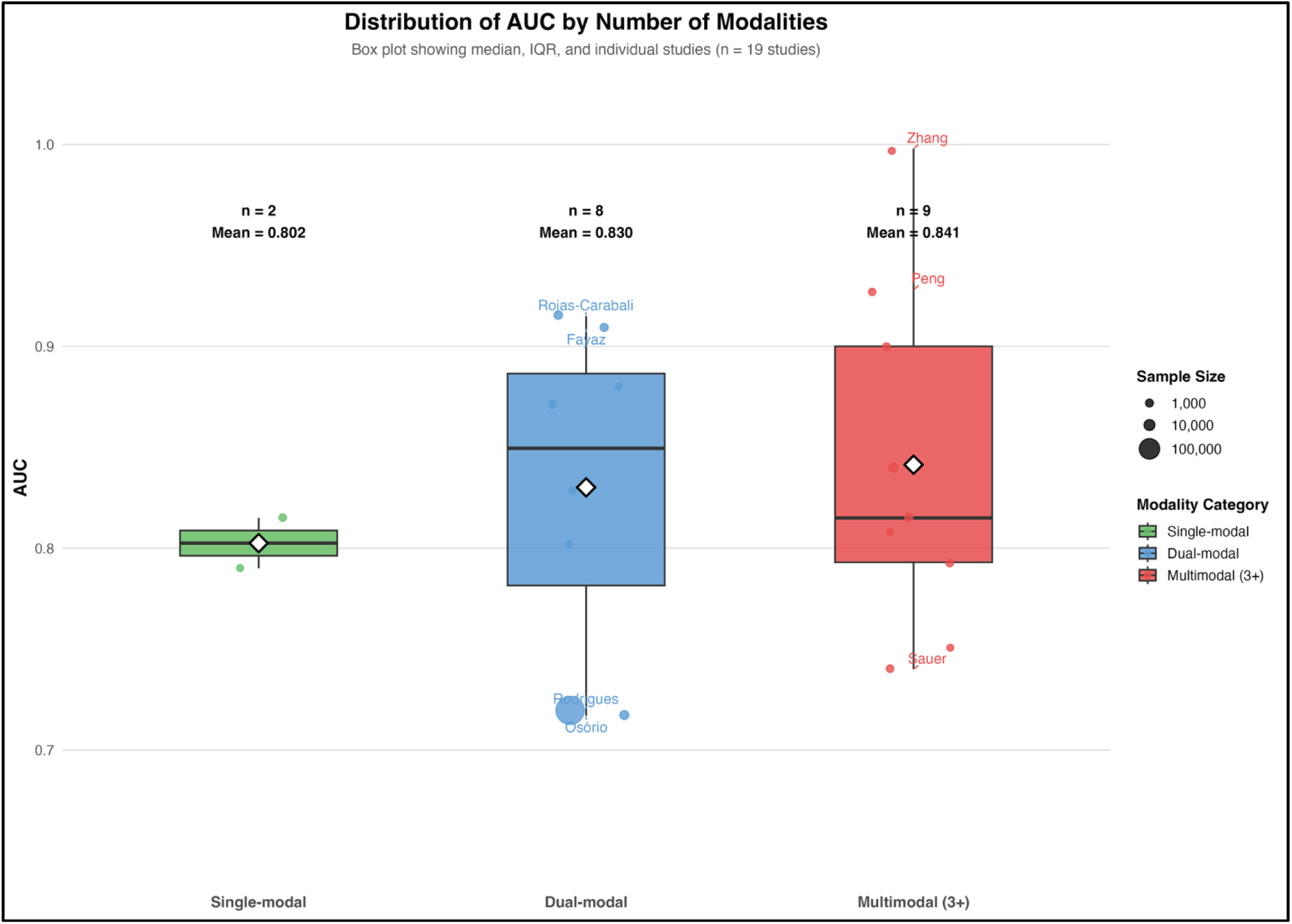
Distribution of Area Under the Curve by Number of Modalities. Box plot comparing area under the curve values across studies grouped by the number of data modalities used: single-modal (n=2 studies), dual-modal (n=8), and multimodal with three or more data types (n=9). Boxes represent the interquartile range, horizontal lines indicate the median, and whiskers extend to the most extreme data points within 1.5 times the interquartile range. Individual study points are overlaid, with point size proportional to sample size. Mean area under the curve increased from 0.802 (single-modal) to 0.830 (dual-modal) to 0.841 (multimodal), but the overall difference was not statistically significant (Kruskal–Wallis p = 0.758). Selected studies are labeled for reference.

However, substantial variability remained within each modality category, indicating that the quality and clinical relevance of predictors may be more important than the number of data domains included. Collectively, these findings suggest that model developers increasingly recognize tuberculosis treatment failure as a multifactorial outcome requiring integration of host, disease, and treatment-related signals rather than reliance on a single predictor domain.

#### Feature selection, explainability, and tuning

Feature selection was reported in 23 studies (67.6%). The most common approaches were filter methods (e.g., univariate statistical tests, mutual information, chi-square; 9 studies), followed by wrapper methods such as recursive feature elimination or stepwise selection (7 studies), and embedded methods including LASSO regularization or random forest importance (6 studies). The Boruta algorithm was used in one study.

Hyperparameter tuning was described in 20 studies (58.8%), most commonly using grid search combined with cross-validation. Model explainability was reported in 12 studies (35.3%), most frequently through feature importance rankings, followed by SHAP values, decision tree visualizations, and odds ratios derived from logistic regression models.

Although feature selection and tuning were commonly reported, explainability methods were inconsistently applied, limiting transparency and interpretability of model predictions for clinical audiences. In addition, incomplete reporting of feature selection and tuning strategies may affect the reproducibility and robustness of model development, contributing to methodological limitations identified in the risk-of-bias assessment.

### Predictive performance

#### Overall pooled discrimination

Nineteen studies contributed AUC data to the meta-analysis, representing 100,790 participants. Study-level discrimination ranged from 0.717 (95% CI 0.677–0.757) in a logistic regression model from Mozambique to 0.998 (95% CI not estimable) in a random forest model from China. The pooled AUC from a random-effects model was 0.836 (95% CI 0.799–0.868), indicating overall good discriminative performance (Figure 7).

**Figure 7.**
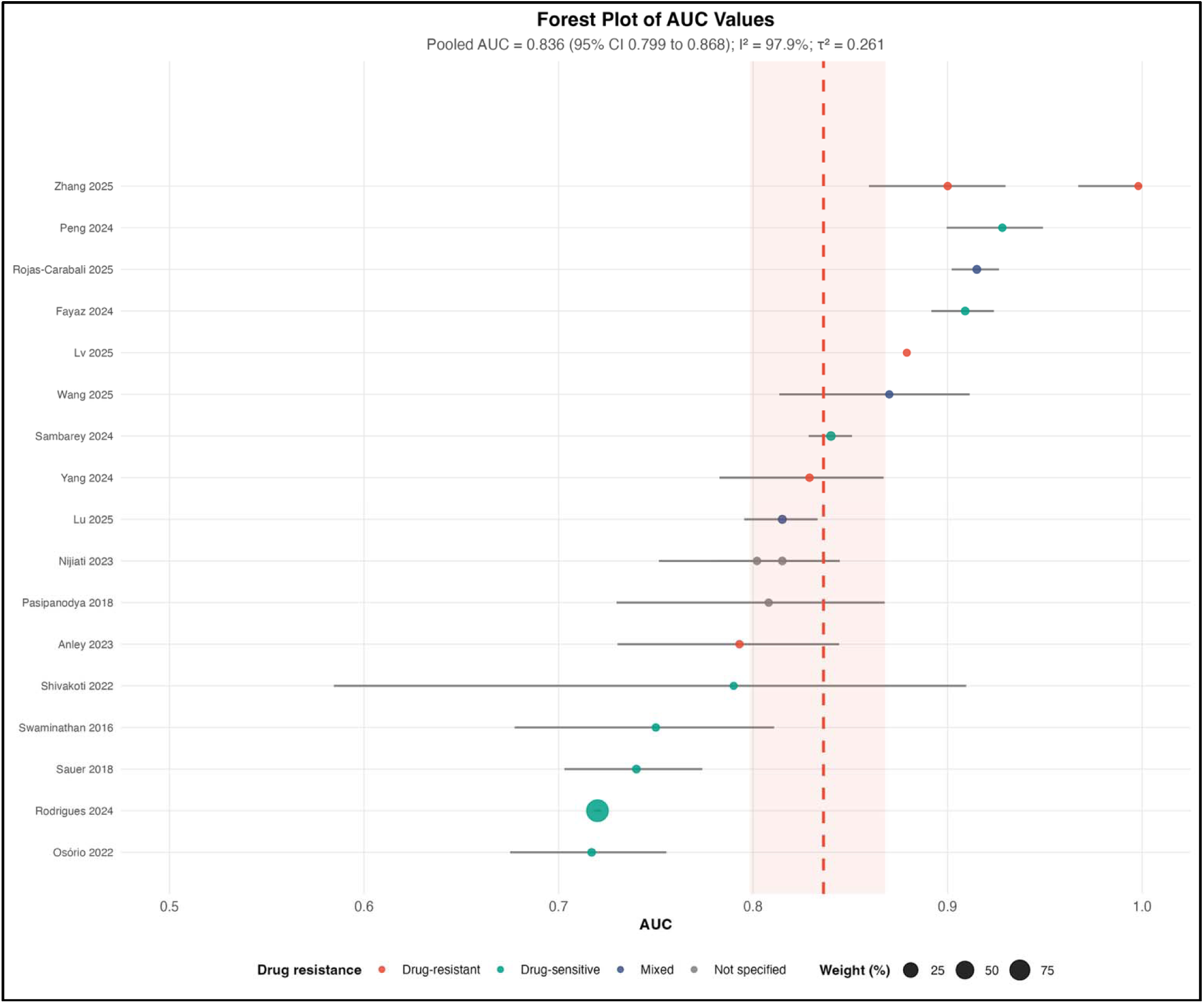
Forest Plot of Area Under the Curve Values. Forest plot displaying individual study area under the curve estimates with 95% confidence intervals for the 19 studies included in the meta-analysis. Squares represent individual study estimates, with square size proportional to the study’s weight in the random-effects meta-analysis. Horizontal lines indicate 95% confidence intervals. The vertical dashed line at area under the curve = 0.5 represents chance performance (no discrimination). The diamond at the bottom represents the pooled random-effects estimate of 0.836 (95% confidence interval 0.799–0.868). Substantial heterogeneity was observed (I² = 97.9%, τ² = 0.261, p < 0.001). Studies are labeled by first author and year; colors indicate drug resistance profile (drug-sensitive, mixed, not specified, or drug-resistant).

However, heterogeneity was extreme (I² = 97.9%, τ² = 0.261, Q p < 0.001), reflecting substantial variability in study populations, predictors, outcome definitions, and modeling approaches. Consequently, the pooled AUC should be interpreted as a summary measure of central tendency rather than a generalizable performance benchmark.

Importantly, discrimination alone does not ensure clinical utility. Models may distinguish higher- from lower-risk individuals while remaining poorly calibrated, non-transportable, or impractical for implementation. In tuberculosis, where outcomes are shaped by microbiological factors, drug resistance, treatment adherence, comorbidities, and health system constraints, predictive performance must be interpreted within this broader clinical and contextual framework.

#### Subgroup analyses

We conducted prespecified subgroup analyses to explore sources of heterogeneity (Table 5; Figure 8). Model performance varied across clinical and methodological strata. Discrimination was higher in drug-resistant tuberculosis (AUC 0.879) and mixed-resistance populations (0.872) than in drug-sensitive cohorts (0.815). Performance differed markedly by HIV status. Models developed in populations including HIV-positive individuals showed substantially lower discrimination (AUC 0.748) compared with those that excluded HIV-positive participants (0.924), indicating limited generalizability across HIV contexts.

**Figure 8.**
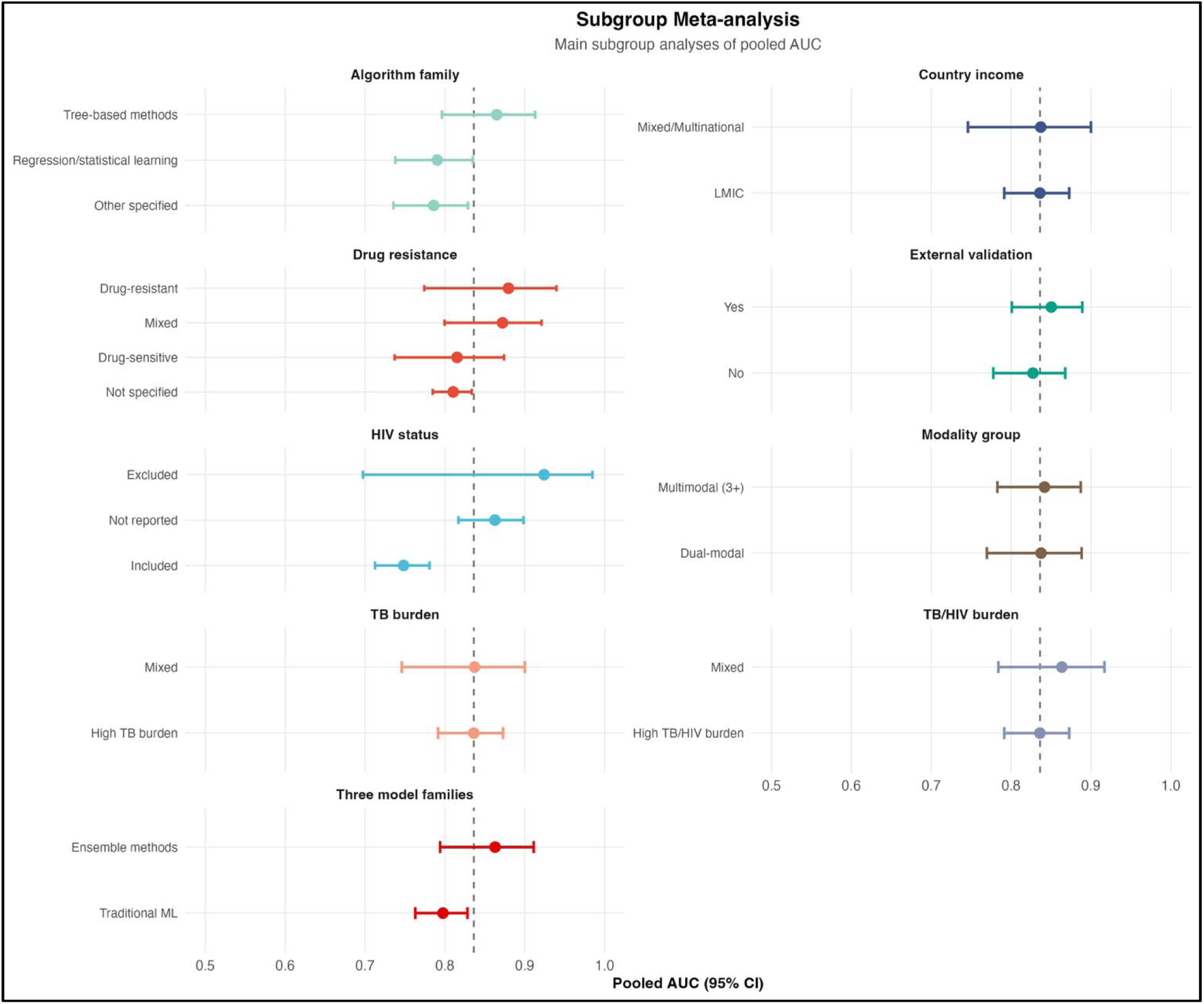
Subgroup Meta-Analysis of Pooled Area Under the Curve. Forest plot displaying pooled area under the curve estimates with 95% confidence intervals for all prespecified subgroup analyses. Subgroups are organized by category: drug resistance profile, HIV status, external validation, country income, tuberculosis burden, tuberculosis/HIV burden, algorithm family, broad algorithm family (traditional machine learning versus ensemble methods), and data modality group. The vertical dashed line at area under the curve = 0.836 indicates the overall pooled estimate. Notable findings include lower pooled area under the curve in studies including HIV-positive participants (0.748) compared to those excluding them (0.924), and higher performance in studies using clinical plus laboratory data (0.910) compared to clinical data alone (0.818).

**Table 5.**
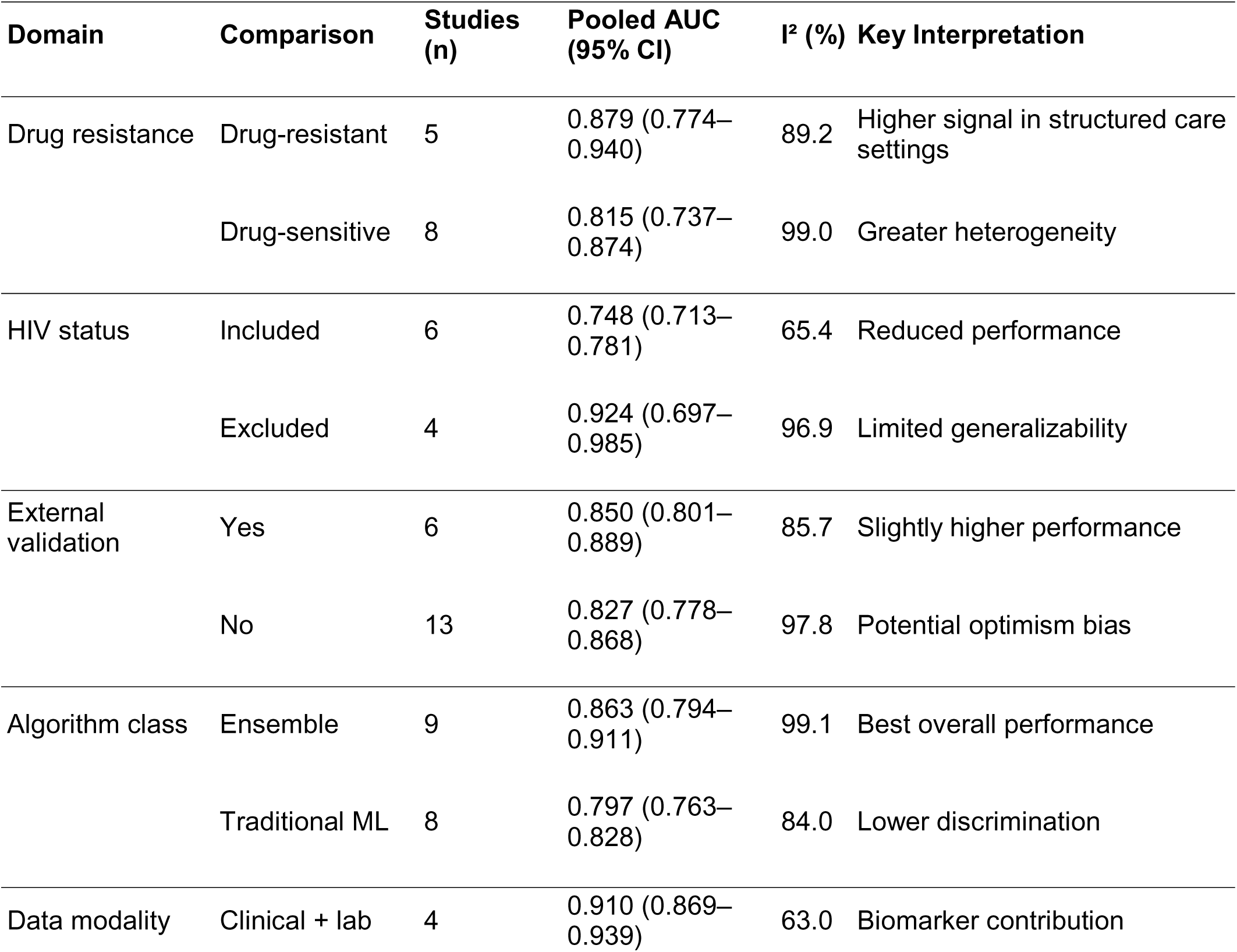
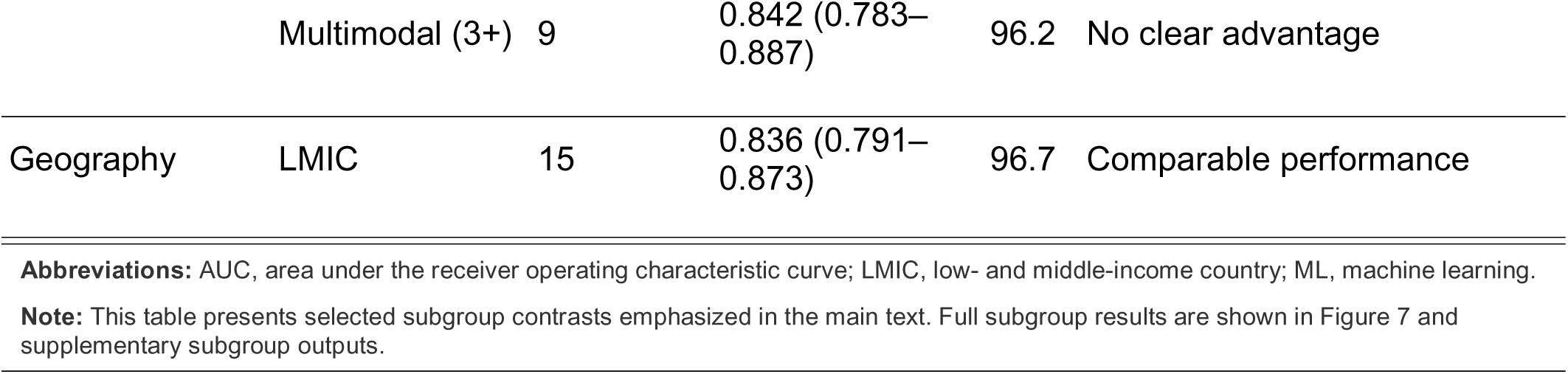
Summary of Subgroup Meta-analysis Findings.

Models with external validation performed similarly to those without (0.850 vs 0.827), although only a minority underwent such validation. By modeling approach, tree-based and ensemble methods outperformed traditional statistical models (AUC ∼0.86 vs ∼0.80), consistent with their ability to capture nonlinear interactions.

Performance differences by data modality were modest. Models incorporating laboratory data showed the highest AUCs (∼0.91), whereas increasing the number of modalities did not significantly improve performance (p = 0.758), suggesting that data relevance outweighs data quantity.

Across geographic and epidemiologic strata, including LMICs, high TB burden, and TB/HIV burden settings, pooled AUCs were similar (∼0.836), though heterogeneity remained high and representation uneven. Sensitivity (31–92%) and specificity (60–96%) varied widely, reflecting differences in prediction thresholds and clinical priorities.

### Meta-regression

We performed meta-regression to examine whether study-level characteristics were associated with model discrimination, measured as logit-transformed AUC (Table 6). In univariable analyses, year of publication was positively associated with performance (β = 0.110, 95% CI 0.026 to 0.194; p = 0.010), indicating modest improvement in reported discrimination over time. Inclusion of HIV-positive participants was associated with lower performance (β = −0.690, 95% CI −1.135 to −0.245; p = 0.002), consistent with subgroup findings. In contrast, sample size, external validation, drug-resistant tuberculosis focus, and high TB burden setting were not significantly associated with AUC. LMIC setting was not estimable, as all studies contributing AUC data were conducted in LMIC settings.

**Table 6.** Meta-regression of study-level factors associated with model performance. Associations with logit-transformed AUC

| Moderator | $\beta$ coefficient | 95% CI | P value | Note |
| --- | --- | --- | --- | --- |
| **Multivariate** |  |  |  |  |
| Year of publication (centered at 2015) | 0.125 | 0.010 to 0.241 | 0.033 | NA |
| Sample size (log-transformed) | -0.136 | -0.289 to 0.016 | 0.080 | NA |
| External validation | -0.161 | -0.659 to 0.336 | 0.525 | NA |
| HIV-positive participants included | -0.331 | -0.923 to 0.262 | 0.274 | NA |
| Drug-resistant tuberculosis focus | -0.034 | -0.616 to 0.549 | 0.910 | NA |
| **Univariate** |  |  |  |  |
| Year of publication (centered at 2015) | 0.110 | 0.026 to 0.194 | 0.010 | NA |
| Sample size (log-transformed) | -0.068 | -0.218 to 0.081 | 0.371 | NA |
| External validation | 0.255 | -0.343 to 0.854 | 0.403 | NA |
| HIV-positive participants included | -0.690 | -1.135 to -0.245 | 0.002 | NA |
| Drug-resistant tuberculosis focus | 0.326 | -0.339 to 0.991 | 0.337 | NA |
| High TB burden setting | -0.007 | -0.620 to 0.607 | 0.983 | NA |
| LMIC setting | , | , | , | NA |

**Abbreviations:** AUC, area under the receiver operating characteristic curve; LMIC, low- and middle-income country.
**Note:** Positive coefficients indicate higher logit-AUC with increasing values of the moderator. The multivariable model adjusted for year, sample size, external validation, HIV inclusion, and drug-resistant tuberculosis focus.

In the multivariable model, adjusting for year, sample size, external validation, HIV inclusion, and drug-resistant tuberculosis focus, year of publication remained independently associated with higher performance (β = 0.125, 95% CI 0.010 to 0.241; p = 0.033). Sample size showed a borderline inverse association (β = −0.137, 95% CI −0.289 to 0.016; p = 0.080), suggesting a tendency for smaller studies to report higher performance. The association between HIV inclusion and performance was attenuated and no longer statistically significant (β = −0.331, 95% CI −0.923 to 0.262; p = 0.274), while external validation and drug-resistant tuberculosis focus remained non-significant.

The multivariable model explained approximately 43% of between-study heterogeneity (R² = 42.99%), although substantial residual heterogeneity remained (I² = 90.3%; p < 0.001), indicating that additional unmeasured factors contribute to variability in model performance.

### Predictor domains and knowledge gaps

Predictor selection varied across studies, with clinical variables used in 31 studies (91.2%), including age, sex, body mass index, symptoms, and comorbidities (Figure 9). Other domains were less consistently represented: social factors (education, income, alcohol use, smoking) and radiological features (chest X-ray, CT, lesion characteristics) were each used in 13 studies (38.2%); microbiological variables (smear, culture, GeneXpert, drug susceptibility testing) in 9 (26.5%); and laboratory markers (blood tests, inflammatory and biochemical parameters) in 7 (20.6%). Genomic or omics features (e.g., DNA methylation, transcriptomics) were included in 4 studies (11.8%), and pharmacokinetic variables (e.g., drug concentrations, Cmax) in 3 studies (8.8%).

**Figure 9.**
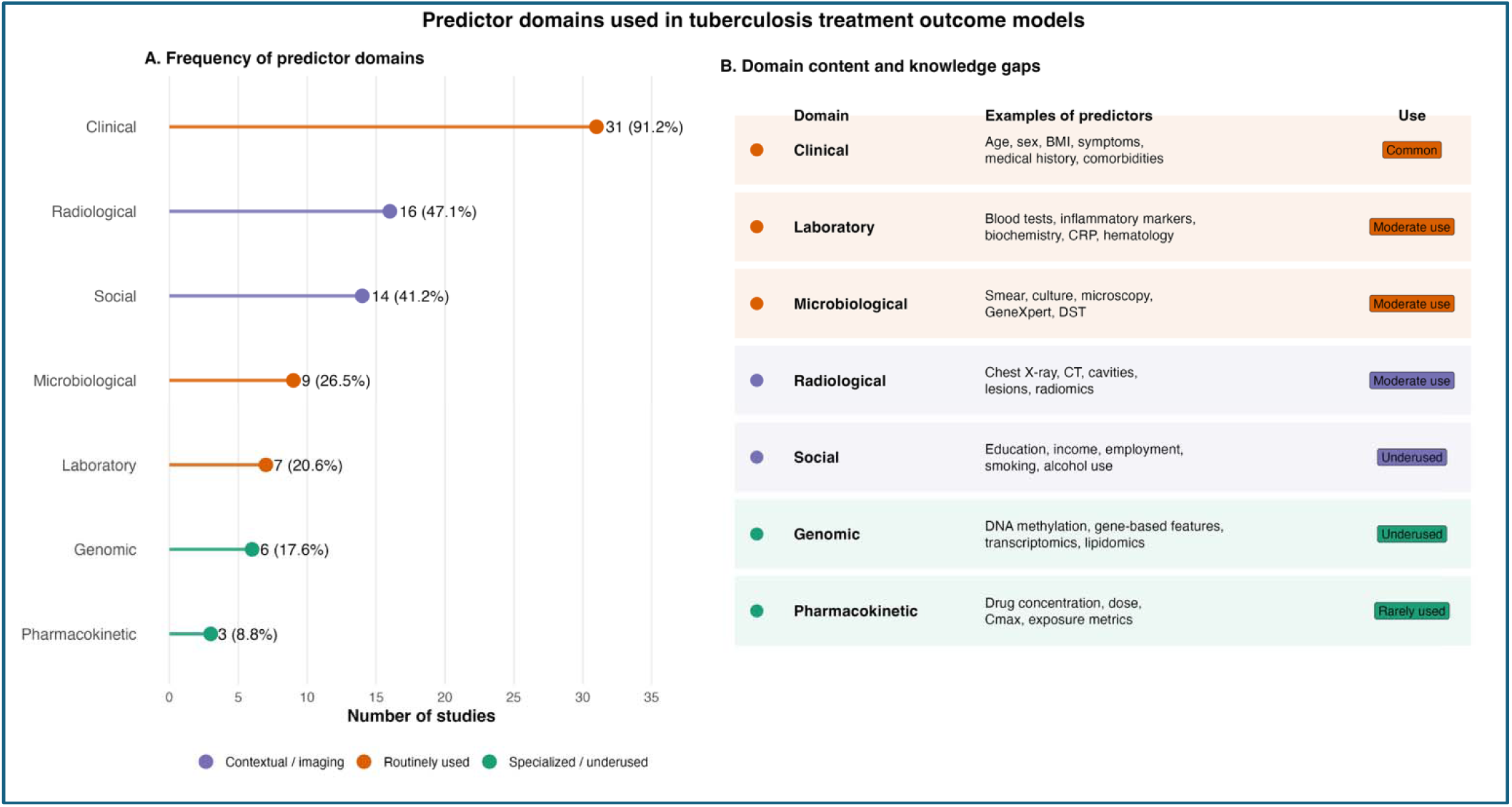
Predictor Domains Used in Tuberculosis Treatment Outcome Models. Two-panel figure summarizing predictor domains across the 34 included studies. **Panel A: Frequency of Predictor Domains.** Bar chart showing the number and percentage of studies using each predictor domain. Clinical and demogr phic variables were most common (31 studies, 91.2%), followed by social determinants (13, 38.2%) and radiological features (13, 38.2%). Microbiological data (9, 26.5%), laboratory biomarkers (7, 20.6%), genomic/omics data (4, 11.8%), and pharmacokinetic data (3, 8.8%) were used less frequently. **Panel B: Domain Content and Knowledge Gaps.** Table summarizing examples of predictors within each domain and their frequency of use. Social determinants, genomic/omics data, and pharmacokinetic data are identified as underused or rarely sed, representing important knowledge gaps in the literature.

Overall, models relied predominantly on routinely available clinical data, with limited incorporation of pharmacologic, molecular, and behavioral predictors. Pharmacokinetic and omics-based features were particularly underrepresented despite their biological relevance, and social or adherence-related variables were inconsistently included. Predictor definitions and measurement approaches were heterogeneous, and few studies incorporated longitudinal data. These findings indicate that current models capture only a partial representation of factors influencing treatment outcomes.

### Methodological quality, validation rigor, and publication bias

#### Risk of bias

Risk of bias was predominantly driven by deficiencies in the analysis domain (Figure 10; Table 7). Although most studies were judged to be at low risk in the participants (94.1%), predictors (100%), and outcome (100%) domains, only one study was rated as low risk overall, with the remainder classified as high risk due to analytical limitations. Study-level assessments (S2) indicate that these limitations were consistent across studies and largely attributable to inappropriate handling of missing data, absence of calibration assessment, suboptimal feature selection strategies, and inadequate validation procedures. These issues have direct implications for model reliability, as discrimination metrics such as AUC may remain high despite poor calibration or overfitting, thereby limiting clinical applicability.

**Figure 10.**
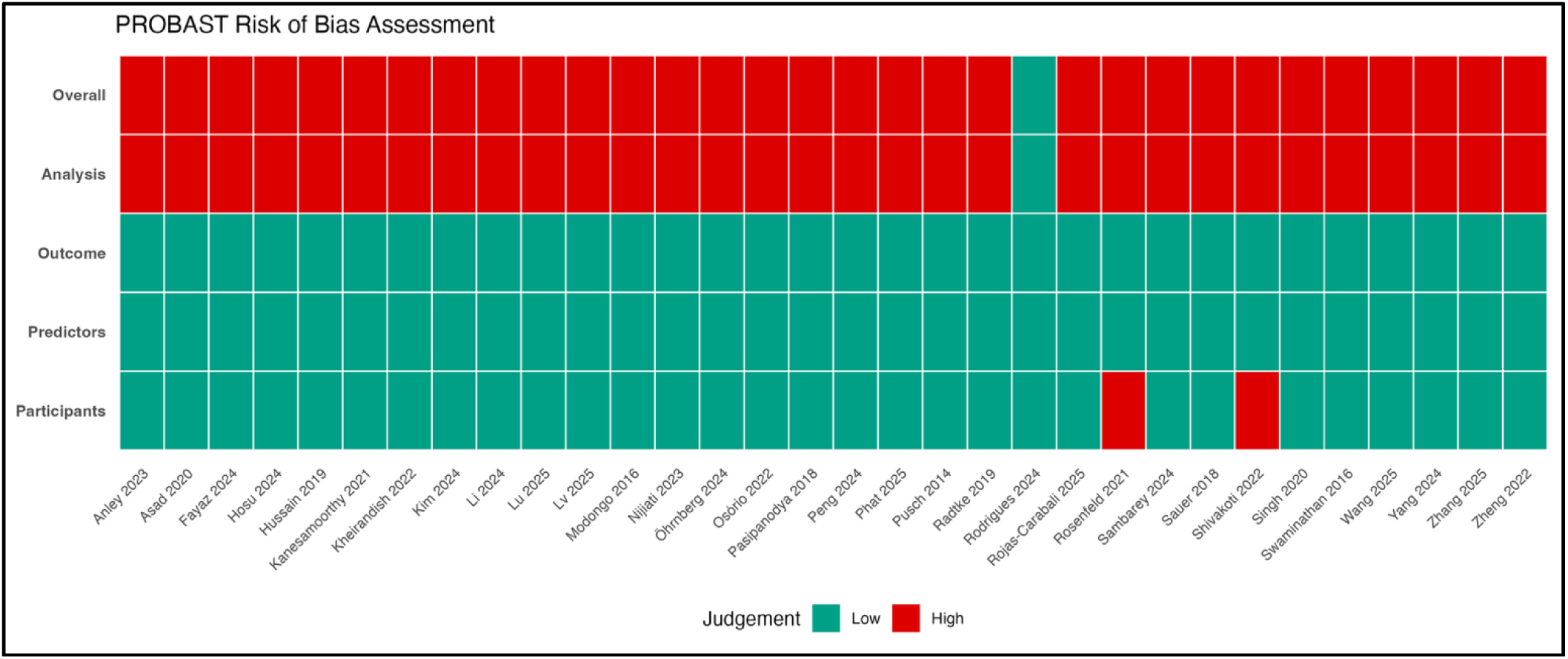
PROBAST Risk of Bias Assessment. Traffic light plot summarizing the risk of bias assessment using the Prediction model Risk Of Bias Assessment Tool (PROBAST). Each row represents one of the 34 included studies, and columns represent the four PROBAST domains (Participants, Predictors, Outcome, Analysis) plus the Overall rating. Green indicates low risk of bias, yellow indicates unclear risk, and red indicates high risk of bias. Only one study (2.9%) was rated as low risk of bias overall. The Analysis domain was the primary source of bias, with 33 studies (97.1%) rated as high risk. The Participants, Predictors, and Outcome domains were predominantly low risk.

**Table 7.**
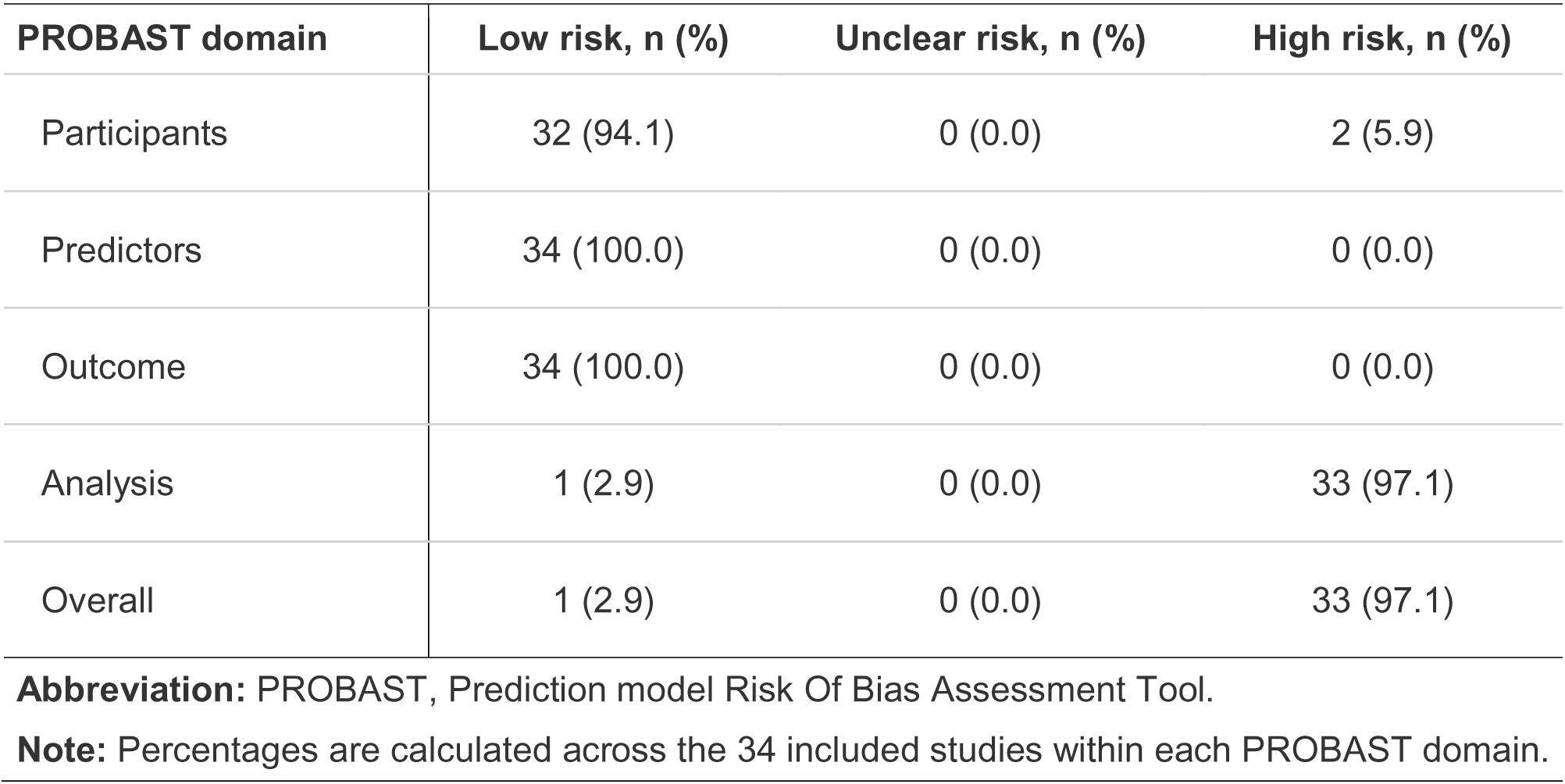
Summary of risk of bias across PROBAST domains. Domain-level distribution of low, unclear, and high risk of bias judgments

| Summary of risk of bias across PROBAST domains |  |  |  |
| --- | --- | --- | --- |
| Domain-level distribution of low, unclear, and high risk of bias judgments |  |  |  |
| PROBAST domain | Low risk, n (%) | Unclear risk, n (%) | High risk, n (%) |

| PROBAST domain | Low risk, n (%) | Unclear risk, n (%) | High risk, n (%) |
| --- | --- | --- | --- |
| Participants | 32 (94.1) | 0 (0.0) | 2 (5.9) |
| Predictors | 34 (100.0) | 0 (0.0) | 0 (0.0) |
| Outcome | 34 (100.0) | 0 (0.0) | 0 (0.0) |
| Analysis | 1 (2.9) | 0 (0.0) | 33 (97.1) |
| Overall | 1 (2.9) | 0 (0.0) | 33 (97.1) |

**Abbreviation:** PROBAST, Prediction model Risk Of Bias Assessment Tool.
**Note:** Percentages are calculated across the 34 included studies within each PROBAST domain.

#### Validation rigor

Most studies reported some form of internal validation (88.2%), primarily using split-sample approaches or k-fold cross-validation; however, external validation was performed in fewer than one-quarter of studies (23.5%). This imbalance underscores a critical limitation, as internal validation alone does not ensure generalizability across settings. Among externally validated models, validation strategies varied in scope, including geographic, temporal, and cross-cohort validation, but the small number of such studies limits robust conclusions regarding transportability.

#### Publication bias

Publication bias was assessed using funnel plot asymmetry and Egger’s test (Figure 11). Visual inspection suggested asymmetry, with fewer small studies reporting lower AUC values. Egger’s test was statistically significant (p = 0.024), indicating small-study effects. The trim-and-fill method did not impute additional studies, suggesting limited impact on the pooled estimate; however, the small number of studies limits interpretability.

**Figure 11.**
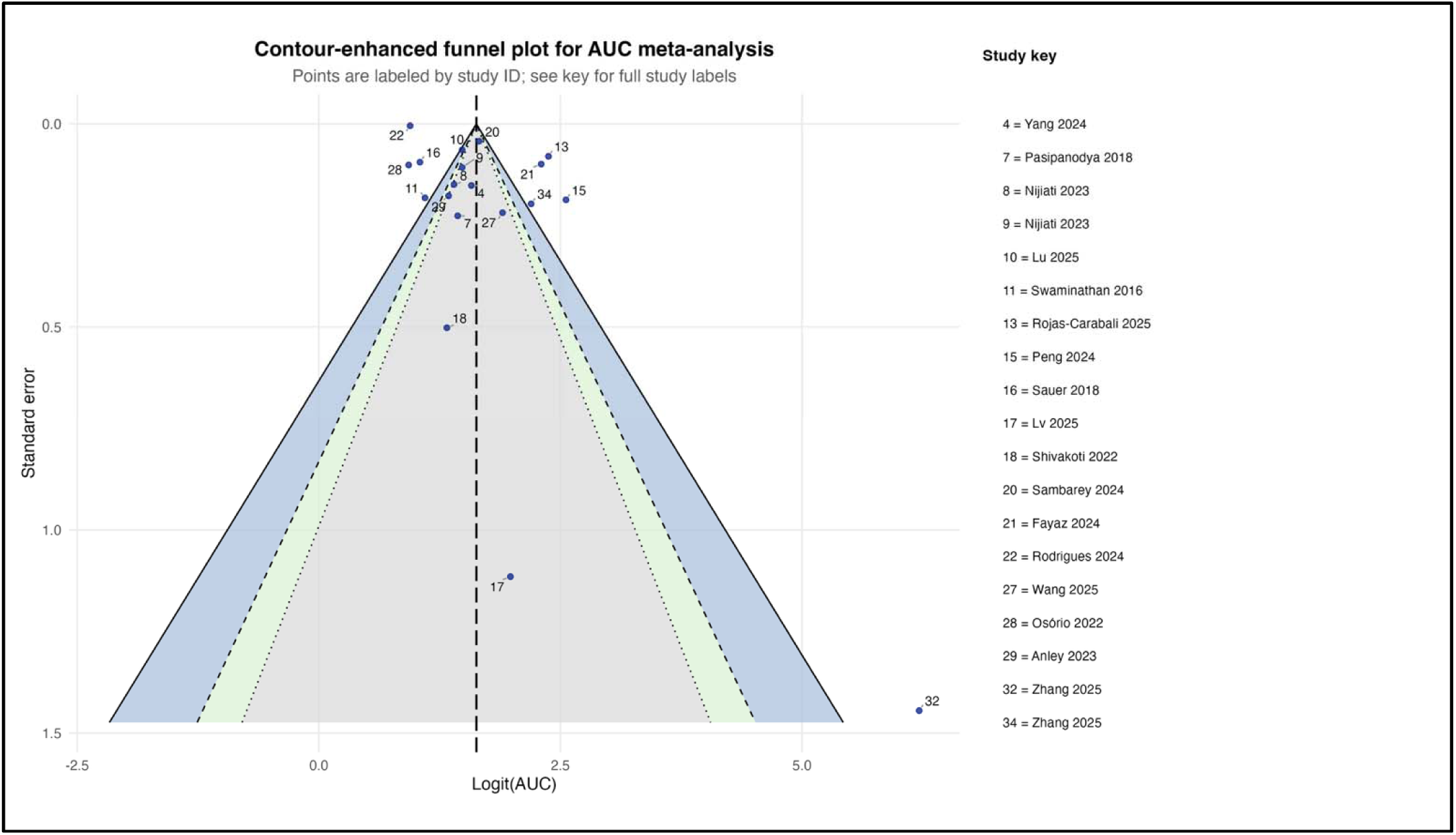
Contour-Enhanced Funnel Plot for Publication Bias Assessment. Contour-enhanced funnel plot for assessing publication bias among the 19 studies included in the area under the curve meta-analysis. Each point represents an individual study, plotted by its logit-transformed area under the curve (x-axis) against its standard error (y-axis). The vertical dashed line represents the pooled estimate. The triangular region represents the expected distribution of studies in the absence of publication bias or small-study effects. Colored contour regions (white, light gray, dark gray) indicate different levels of statistical significance for hypothetical missing studies. Points are labeled by study identification number; a key mapping identification numbers to author names and years is provided. Visual inspection suggests some asymmetry, with a relative absence of small studies (high standard error) reporting low area under the curve values. Egger’s test was significant (p = 0.024), indicating evidence of small-study effects or publication bias.

### Evidence gaps

This review identified several critical gaps that limit the generalizability, equity, and clinical utility of current prediction models.

#### Geographic representation

Despite tuberculosis being concentrated in low- and middle-income countries, most studies were conducted in a small number of settings, primarily China and India. High-burden regions in sub-Saharan Africa and Southeast Asia were sparsely represented, raising concerns about model transportability across diverse epidemiological and health system contexts.

#### HIV-affected populations

Only a minority of studies included HIV-positive participants, and model performance was consistently lower in these populations. Given the substantial burden of TB–HIV co-infection, particularly in sub-Saharan Africa, this represents a major gap requiring dedicated model development and validation.

#### Social determinants

Fewer than 40% of studies incorporated social or behavioral predictors, despite their well-established role in tuberculosis outcomes. Omission of these variables may limit both predictive performance and the identification of actionable intervention targets.

#### Pediatric and extrapulmonary tuberculosis

No studies focused exclusively on children or extrapulmonary disease, and these groups were underrepresented overall. Both populations have distinct clinical and prognostic profiles, necessitating tailored modeling approaches.

#### Validation and calibration

External validation was uncommon, and calibration was rarely assessed. This is a critical limitation, as models with good discrimination may still provide inaccurate risk estimates, undermining clinical decision-making and safe implementation.

#### Implementation and health system factors

No studies evaluated real-world impact or incorporated health system variables such as access to care, treatment support, or drug supply continuity. These factors are central to outcomes in programmatic settings and are essential for models intended for clinical use.

Collectively, these gaps indicate that current models provide an incomplete and context-limited representation of treatment risk. Future work should prioritize geographically diverse datasets, inclusion of HIV-affected and underrepresented populations, integration of social and health system determinants, and rigorous external validation with calibration and implementation evaluation.

## Discussion

This systematic review and meta-analysis synthesizes evidence on artificial intelligence and machine learning models for predicting tuberculosis treatment failure. Across 34 studies and nearly 1.1 million participants, models demonstrated good overall discrimination (pooled AUC 0.836; Figure 7), but this signal was accompanied by extreme heterogeneity (I² = 97.9%), limited external validation (23.5%; Table 7), pervasive high risk of bias (97.1% high risk overall; Table 7; Figure 10), and evidence of small-study effects (Figure 11). Importantly, the geographic distribution of studies was highly uneven (Table 1; Figure 4-5), with limited representation of low- and middle-income countries (LMICs) bearing a dual high burden of tuberculosis and HIV [1,8]. Taken together, these findings indicate that while AI/ML approaches are promising, current models remain insufficiently robust and inadequately representative for routine clinical implementation.

The pooled AUC of 0.836 indicates good discrimination and is comparable to performance reported in prognostic models across infectious diseases and clinical medicine [13,37]. Similar performance has been observed in machine learning applications in healthcare more broadly [15–17]. However, discrimination alone is insufficient for clinical utility, as emphasized in prediction model methodology literature [37,39]. Models may distinguish higher- from lower-risk patients while remaining poorly calibrated or non-transportable, limiting their usefulness in practice [39,40]. In tuberculosis, this limitation is particularly relevant given that treatment outcomes are shaped by microbiological, behavioral, and health system factors rather than risk prediction alone [5,10].

The extreme heterogeneity observed is consistent with prior systematic reviews of prediction models, where variability in outcome definitions and study design is a major driver of between-study differences [28,29]. In this review, outcome definitions varied substantially (Table 4), ranging from WHO-aligned bacteriological failure to composite unfavorable outcomes. Similar inconsistencies have been reported in tuberculosis outcome studies [6,7], limiting comparability and contributing to heterogeneity in pooled estimates.

Model performance was substantially lower in studies including HIV-positive populations (Table 5), consistent with established evidence that TB–HIV co-infection introduces significant clinical complexity [8,44]. HIV alters immune responses, disease presentation, and treatment outcomes, complicating prediction [44,45]. Given that tuberculosis remains a leading cause of mortality among people living with HIV and is concentrated in high-burden LMIC settings [1,8], the underrepresentation of these populations in model development is a critical limitation. This gap has also been highlighted in broader tuberculosis outcome research [26].

Higher discrimination observed in drug-resistant tuberculosis cohorts (Table 5) may reflect more structured care pathways, higher event rates, and closer clinical monitoring. Similar patterns have been reported in tuberculosis treatment outcome studies and meta-analyses of multidrug-resistant disease [6,7]. These structured settings may enhance signal detection compared to drug-susceptible tuberculosis, where outcomes are influenced by more heterogeneous factors, including adherence and social determinants [5].

The predominance of clinical predictors and limited incorporation of molecular, pharmacokinetic, and social variables (Figure 9) mirrors broader trends in clinical machine learning, where models often rely on readily available data rather than biologically rich or contextually relevant predictors [19,46]. While multimodal models showed numerically higher performance (Figure 6), differences were not statistically significant, consistent with evidence that predictor quality is more important than data volume [47]. Notably, social determinants, well-established drivers of tuberculosis outcomes [10,11], were underrepresented, suggesting that current models may inadequately capture structural risk factors.

The dominance of tree-based and ensemble methods is consistent with the broader machine learning literature, where these approaches perform well in complex, nonlinear datasets [15,48,49]. Their flexibility allows modeling of interactions and heterogeneous predictors but also increases the risk of overfitting when validation is insufficient [50]. This is particularly relevant given the high heterogeneity and limited external validation observed in this review.

The high prevalence of analytical bias (Table 7; Figure 10) aligns with findings from previous evaluations of prediction models using PROBAST [22,23]. Common issues, including inappropriate handling of missing data, lack of calibration assessment, and inadequate validation, are well-recognized threats to model validity [52,53]. The absence of calibration reporting is particularly concerning, as calibration is essential for translating model predictions into clinical decision-making [39,54].

External validation was performed in fewer than one-quarter of studies (Table 7), consistent with broader evidence that many prediction models are not adequately validated before publication [24,57]. Internal validation alone cannot ensure generalizability, particularly in tuberculosis, where epidemiological and health system differences across settings are substantial [24]. Methodological guidance emphasizes that external validation is a minimum requirement for clinical implementation [13,24].

The uneven geographic distribution of studies (Table 1; Figure 4-5) reflects broader disparities in global health research [1]. Many high-burden countries, particularly in sub-Saharan Africa, were underrepresented despite bearing the greatest burden of TB and TB–HIV co-infection [1,8]. This imbalance raises concerns about model transportability, as differences in population characteristics, healthcare systems, and disease epidemiology can substantially affect performance.

The limited inclusion of social determinants and health system variables contrasts with extensive evidence demonstrating their central role in tuberculosis outcomes [5,10,11,60]. Factors such as poverty, food insecurity, and access to care strongly influence treatment adherence and success. Their omission in most models suggests a disconnect between predictive modeling and real-world determinants of disease outcomes, potentially limiting clinical relevance.

Funnel plot asymmetry and a significant Egger’s test (Figure 11) suggest the presence of small-study effects, consistent with broader concerns about publication bias in medical research and machine learning [35,63,64]. Although trim-and-fill analysis did not materially alter pooled estimates, the possibility of optimism bias remains, particularly in emerging computational fields [65].

This review extends prior work on machine learning in tuberculosis, which has largely focused on diagnostic applications or narrative summaries [66–68]. By contrast, this study provides quantitative synthesis of predictive performance alongside systematic assessment of heterogeneity and methodological quality. The pooled AUC observed here is consistent with recent meta-analyses of machine learning models for TB outcomes [69], but the extreme heterogeneity highlights the need for more standardized and rigorous research.

For clinicians, these findings support cautious interpretation of existing models, particularly in settings that differ from development populations. For researchers, priorities include improving methodological rigor, incorporating underrepresented predictor domains, and focusing on high-burden populations, particularly those affected by HIV. For policymakers, investment should extend beyond model development to include validation infrastructure, data sharing, and implementation research, particularly in LMIC settings where the burden of disease is greatest [71–73].

### Strengths and limitations

This study has several strengths. First, it provides a comprehensive and up-to-date synthesis of artificial intelligence and machine learning models for predicting tuberculosis treatment failure, incorporating both qualitative and quantitative evidence. By including 34 studies spanning diverse geographic regions, clinical settings, and populations, this review captures the breadth of current modeling approaches and their real-world variability. Second, the study integrates meta-analysis of discrimination with detailed evaluation of methodological quality, predictor domains, and validation practices, offering a multidimensional assessment rather than focusing solely on performance metrics.

Third, the use of established methodological frameworks, including PRISMA for reporting [27], PROBAST for risk-of-bias assessment [22,23], and recommended approaches for meta-analysis of prediction models [28,29], enhances transparency, reproducibility, and rigor. Fourth, the incorporation of subgroup analyses and meta-regression provides insights into sources of heterogeneity, including the impact of HIV status, drug resistance, and study design on model performance. Finally, the study explicitly examines evidence gaps, including geographic representation, predictor selection, and validation, thereby informing priorities for future research.

Several limitations should be considered. First, only a subset of included studies reported sufficient data for meta-analysis, restricting quantitative synthesis to 19 studies and potentially introducing selection bias. Second, heterogeneity across studies was substantial, reflecting differences in populations, predictor sets, outcome definitions, and modeling approaches; although expected in this field, it limits the interpretability of pooled estimates. Third, outcome definitions for “treatment failure” were inconsistent (Table 4), which may have contributed to variability in performance and complicates direct comparison across studies. Fourth, most studies were rated as high risk of bias, particularly in the analysis domain (Table 7; Figure 10), which limits confidence in reported performance estimates. Fifth, important aspects of model evaluation, especially calibration and external validation, were poorly reported, constraining assessment of clinical utility. Sixth, the geographic distribution of studies was uneven, with limited representation from high TB/HIV burden settings in sub-Saharan Africa (Table 1; Figure 3), restricting generalizability. Seventh, the review was limited to English-language publications, which may have excluded relevant studies. Finally, although publication bias was assessed using established methods [35,36], the relatively small number of studies limits the robustness of these analyses.

## Conclusion

Machine learning models for predicting tuberculosis treatment failure demonstrate promising discriminative performance, with a pooled AUC of 0.836, but are constrained by substantial heterogeneity, high risk of bias, limited external validation, and gaps in population and predictor representation. In particular, the underrepresentation of high-burden LMIC settings and HIV-affected populations raises concerns about the generalizability and equity of current models. Moreover, the limited incorporation of social, pharmacologic, and molecular predictors suggests that existing models capture only a partial representation of the determinants of treatment outcomes.

Advancing the field will require a shift from model development toward methodological rigor and clinical translation. Future studies should prioritize standardized outcome definitions, robust handling of missing data, routine assessment of calibration, and rigorous external validation across diverse settings. Greater inclusion of underrepresented populations, particularly those affected by HIV, as well as integration of social and health system determinants, will be essential for improving both performance and relevance. In addition, implementation-focused research is needed to evaluate whether prediction models improve clinical decision-making and patient outcomes in real-world settings.

Taken together, current evidence supports the potential of artificial intelligence and machine learning in tuberculosis care but underscores that most models remain investigational. Bridging the gap between predictive performance and clinical utility will be critical to realizing the promise of these approaches in high-burden settings.

## Methods

### Study design and registration

This systematic review and meta-analysis was conducted in accordance with the Preferred Reporting Items for Systematic Reviews and Meta-Analyses (PRISMA) 2020 statement and informed by methodological guidance for systematic reviews of prediction model studies and diagnostic accuracy evidence [26,27]. The protocol was prospectively registered in PROSPERO (CRD420251101443).

### Search strategy

We systematically searched PubMed/MEDLINE and Embase for studies published from 1 January 2000 to 31 October 2025. The search combined controlled vocabulary terms, including Medical Subject Headings and Emtree headings, with free-text terms related to tuberculosis, artificial intelligence, machine learning, prediction modeling, and treatment outcomes. Search development was informed by established systematic review principles and structured to maximize sensitivity and reproducibility. The full search strategies are provided in S5.

### Eligibility criteria

Eligibility was defined using a modified Population, Intervention, Comparator, Outcome framework. We included studies of adults or children with active pulmonary or extrapulmonary tuberculosis, including both drug-susceptible and drug-resistant disease, who were undergoing anti-tuberculosis treatment. Eligible studies developed, validated, or implemented an artificial intelligence or machine learning model for prediction of treatment failure or a closely related poor treatment outcome. Because this was a review of prediction modeling studies, a conventional comparator was not required. Studies were required to report at least one quantitative measure of predictive performance, such as area under the curve, sensitivity, specificity, accuracy, or F1-score. We excluded studies that used only conventional statistical methods without a machine learning component; studies focused exclusively on tuberculosis diagnosis, latent tuberculosis infection, or transmission without treatment outcome prediction; studies without original data, including reviews, editorials, and commentaries; animal or in vitro studies; and non-English publications.

### Study selection

Three reviewers independently screened titles and abstracts using Rayyan. Full texts of potentially eligible records were then reviewed independently by the same reviewers against the predefined criteria. Disagreements were resolved through discussion or, when necessary, consultation with a fourth reviewer. The study selection process was documented using a PRISMA flow diagram.

### Data extraction

A standardized extraction form was developed a priori and piloted before full extraction, consistent with guidance for prediction model reviews [27,28]. Extracted data included study design, setting, country, recruitment source, study period, sample size, population characteristics, drug susceptibility profile, HIV status, comorbidities, treatment failure definition, follow-up duration, predictor domains, data modalities, feature engineering and selection methods, algorithms evaluated, model validation strategies, hyperparameter tuning, discrimination and calibration measures, explainability methods, and data required for Prediction model Risk Of Bias Assessment Tool assessment.

### Risk of bias assessment

Methodological quality and risk of bias were assessed using the Prediction model Risk Of Bias Assessment Tool, which evaluates prediction studies across the domains of participants, predictors, outcome, and analysis [22,23]. Each domain was rated as low, high, or unclear risk of bias according to Prediction model Risk Of Bias Assessment Tool guidance, and an overall judgment was derived accordingly.

### Data synthesis and statistical analysis

The primary quantitative outcome was model discrimination, summarized using the area under the curve. Because area under the curve values are bounded between 0 and 1, we applied a logit transformation before meta-analysis to improve statistical behavior and stabilize variance [24,27].

Standard errors were derived from reported confidence intervals when available or estimated using established methods when necessary [24]. Random-effects meta-analysis was performed using restricted maximum likelihood estimation, and pooled estimates were back-transformed to the original area under the curve scale for interpretation [29,30]. Statistical heterogeneity was assessed using Cochran’s Q and the I² statistic [31].

We anticipated substantial heterogeneity given expected differences in study populations, settings, predictors, outcome definitions, and model families. Accordingly, we conducted prespecified subgroup analyses by tuberculosis susceptibility profile, validation status, country income context, and data modality, with particular interest in comparisons across high versus lower tuberculosis burden settings, tuberculosis and HIV relevant populations, and low- and middle-income country versus high-income country contexts where data permitted. For studies reporting both sensitivity and specificity, we planned to use a bivariate random-effects model based on the Reitsma approach to account for the correlation between these measures and to generate summary receiver operating characteristic estimates [32,33], but insufficient studies preformed this analysis.

Meta-regression using mixed-effects models was performed to examine whether study-level covariates, including publication year, sample size, tuberculosis burden setting, external validation, drug resistance profile, HIV inclusion, and model complexity, were associated with area under the curve [27]. Sensitivity analyses included leave-one-out meta-analysis and restriction to studies with lower risk of bias or more rigorous validation. Publication bias was assessed using funnel plot inspection and Egger’s regression test, and where asymmetry was present, we planned to apply the trim-and-fill method to estimate its possible effect on pooled discrimination [34,35].

All analyses were conducted in R version 4.5.2 using the meta, metafor, mada, robvis, and ggplot2 packages [30]. Statistical tests were two-sided, and a p-value less than 0.05 was considered statistically significant.

## Supporting information

PRISMA checklist

Table_S1_study_characteristics

Table_S2_probast_study_level

Search strategy

## Data Availability

All relevant data are within the manuscript and its Supporting Information files

## Acknowledgments

We thank the authors of the primary studies whose work formed the evidence base for this review. We acknowledge the contributions of the screening, data extraction, and analytic team members who supported review conduct and synthesis. We are grateful to colleagues and mentors who provided methodological and domain-specific input during protocol development, risk of bias assessment, and interpretation of findings. Finally, we are grateful for the administrative assistance from Ms. Harriet Nakayiza.

## Author Contributions

**Conceptualization:** David Patrick Kateete, Joyce Nakatumba-Nabende, Florence Nameere Kivunike and Rogers Kamulegeya

**Data curation:** Rogers Kamulegeya, Rose Nabatanzi, Derrick Semugenze, Faridah Mugala, Mercy Takuwa, Emmanuel Nasinghe.

**Formal analysis:** Rogers Kamulegeya, Derrick Semugenze, Denis Musinguzi.

**Methodology:** Rogers Kamulegeya, Derrick Semugenze, Joyce Nakatumba-Nabende, Florence Nameere Kivunike, David Patrick Kateete.

**Supervision:** David Patrick Kateete, Joyce Nakatumba-Nabende, Florence Nameere Kivunike, Willy Ssengooba.

**Writing** – manuscript draft: Rogers Kamulegeya.

**Writing** – review & editing: All authors.

## Supplementary information

**Figure.**
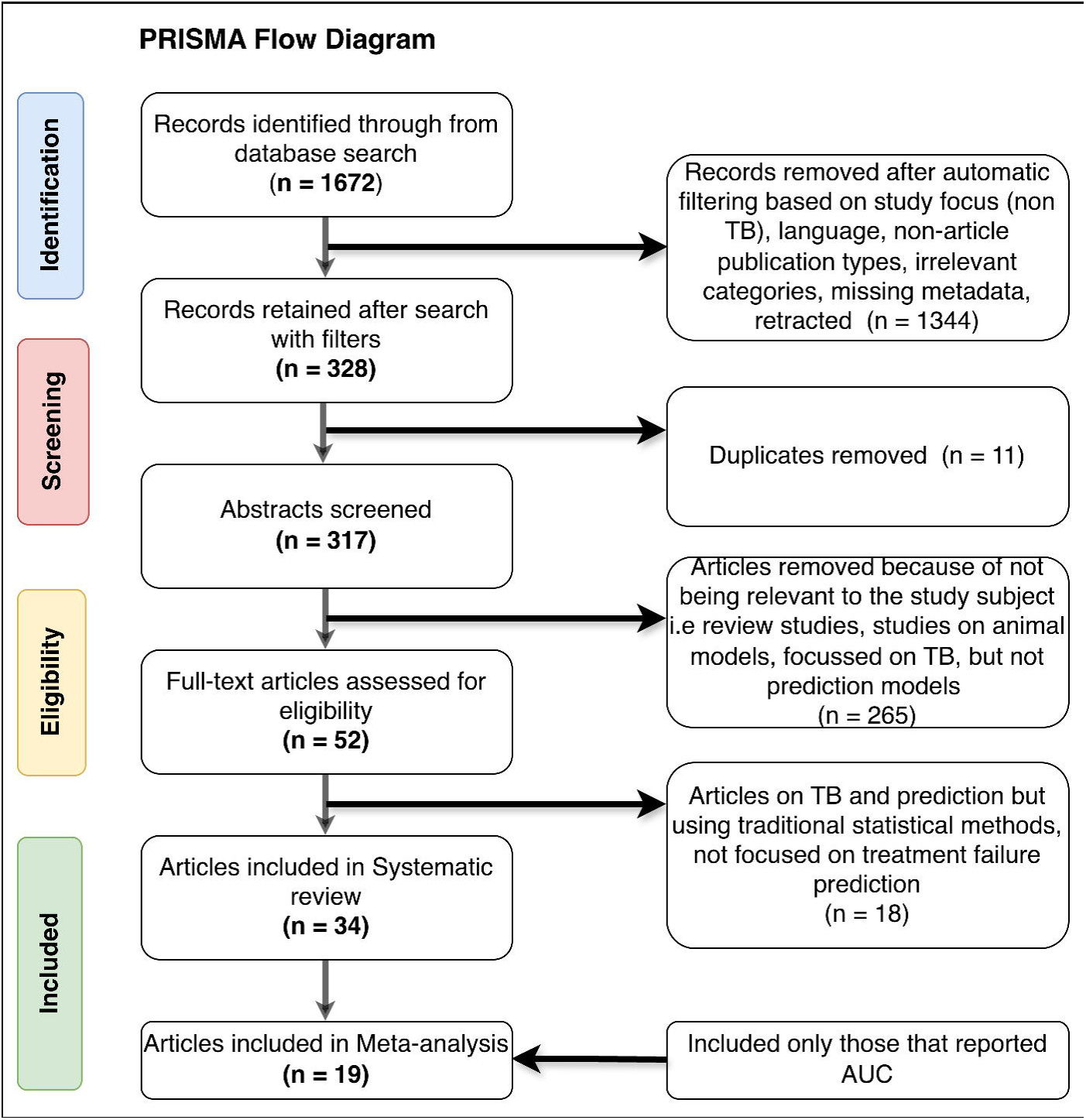
PRISMA Flow Diagram. Flow diagram documenting the study selection process for the systematic review and meta-analysis. From an initial 1,672 records identified through database searching, 1,344 records were removed during initial filtering based on study focus (non-tuberculosis), language, non-article publication types, irrelevant categories, missing metadata, or retraction status. After removal of 11 duplicates, 317 abstracts were screened, of which 265 were excluded as not relevant (review studies, animal models, tuberculosis-focused but not prediction models). Fifty-two full-text articles were assessed for eligibility; 18 were excluded for using traditional statistical methods without a machine learning component or not focusing on treatment failure prediction. Thirty-four studies were included in the systematic review; 19 reported area under the curve values and were included in the meta-analysis.

**Table S1:** Study characteristics

**Table S2:** Probast study level

**Table S3:** Study characteristics, cont’d.

**S4 File:** PROSPERO Registered Protocol

**S5** – Search strategy

