## Supplementary material for "AI/ML-based prediction of TB treatment failure: A systematic review and meta-analysis": PRISMA checklist

### PRISMA 2020 Checklist

| Section and Topic | Item # | Checklist item | Location where item is reported |
|---|---|---|---|
| **TITLE** | | | |
| Title | 1 | Identify the report as a systematic review. | Title – Page 1 |
| **ABSTRACT** | | | |
| Abstract | 2 | See the PRISMA 2020 for Abstracts checklist. | Abstract – Page 2 |
| **INTRODUCTION** | | | |
| Rationale | 3 | Describe the rationale for the review in the context of existing knowledge. | Introduction – Page 4 to 6; line 85 -110 |
| Objectives | 4 | Provide an explicit statement of the objective(s) or question(s) the review addresses. | Introduction– Page 4 to 6; line 114 - 120 |
| **METHODS** | | | |
| Eligibility criteria | 5 | Specify the inclusion and exclusion criteria for the review and how studies were grouped for the syntheses. | Methods: Eligibility criteria – Page 37; Line 627 - 639 |
| Information sources | 6 | Specify all databases, registers, websites, organisations, reference lists and other sources searched or consulted to identify studies. Specify the date when each source was last searched or consulted. | Methods: Search strategy – Page 37; Line 620 - 626 |
| Search strategy | 7 | Present the full search strategies for all databases, registers and websites, including any filters and limits used. | Methods: Search strategy – Page 37; Line 620 - 626 |
| Selection process | 8 | Specify the methods used to decide whether a study met the inclusion criteria of the review, including how many reviewers screened each record and each report retrieved, whether they worked independently, and if applicable, details of automation tools used in the process. | Methods: Selection process – Page 37 to 38; Line 640 - 644 |
| Data collection process | 9 | Specify the methods used to collect data from reports, including how many reviewers collected data from each report, whether they worked independently, any processes for obtaining or confirming data from study investigators, and if applicable, details of automation tools used in the process. | Methods: Data extraction – Page 38; Line 645 - 652 |
| Data items | 10a | List and define all outcomes for which data were sought. Specify whether all results that were compatible with each outcome domain in each study were sought (e.g. for all measures, time points, analyses), and if not, the methods used to decide which results to collect. | Methods: Data extraction – Page 38; Line 645 - 652 |
| | 10b | List and define all other variables for which data were sought (e.g. participant and intervention characteristics, funding sources). Describe any assumptions made about any missing or unclear information. | Methods: Data extraction – Page 38; Line 645 - 652 |
| Study risk of bias assessment | 11 | Specify the methods used to assess risk of bias in the included studies, including details of the tool(s) used, how many reviewers assessed each study and whether they worked independently, and if applicable, details of automation tools used in the process. | Methods: Risk of bias assessment – Page 38; Line 653 - 658 |
| Effect measures | 12 | Specify for each outcome the effect measure(s) (e.g. risk ratio, mean difference) used in the synthesis or presentation of results. | Methods: Data synthesis – Page 38; Line 659 - 685 |
| Synthesis methods | 13a | Describe the processes used to decide which studies were eligible for each synthesis (e.g. tabulating the study intervention characteristics and comparing against the planned groups for each synthesis (item #5)). | Methods: Data synthesis – Page 38; Line 659 - 685 |
| | 13b | Describe any methods required to prepare the data for presentation or synthesis, such as handling of missing summary statistics, or data conversions. | Methods: Data synthesis – Page 38; Line 659 - 685 |
| | 13c | Describe any methods used to tabulate or visually display results of individual studies and syntheses. | Methods: Data synthesis – Page 38; Line 659 - 685 |
| | 13d | Describe any methods used to synthesize results and provide a rationale for the choice(s). If meta-analysis was performed, describe the model(s), method(s) to identify the presence and extent of statistical heterogeneity, and software package(s) used. | Methods: Data synthesis – Page 38; Line 659 - 685 |

### PRISMA 2020 Checklist

| Section and Topic | Item # | Checklist item | Location where item is reported |
|---|---|---|---|
| | 13e | Describe any methods used to explore possible causes of heterogeneity among study results (e.g. subgroup analysis, meta-regression). | Methods: Data synthesis – Page 38; Line 659 - 685 |
| | 13f | Describe any sensitivity analyses conducted to assess robustness of the synthesized results. | Methods: Data synthesis – Page 38; Line 659 - 685 |
| Reporting bias assessment | 14 | Describe any methods used to assess risk of bias due to missing results in a synthesis (arising from reporting biases). | Methods: Data synthesis – Page 38; Line 659 - 685 |
| Certainty assessment | 15 | Describe any methods used to assess certainty (or confidence) in the body of evidence for an outcome. | Methods: Data synthesis – Page 38; Line 659 - 685 |
| **RESULTS** | | | |
| Study selection | 16a | Describe the results of the search and selection process, from the number of records identified in the search to the number of studies included in the review, ideally using a flow diagram. | Results: Study selection – Page 6; Line 122 - 135 |
| | 16b | Cite studies that might appear to meet the inclusion criteria, but which were excluded, and explain why they were excluded. | Results: Study selection – Page 6; Line 122 - 135 |
| Study characteristics | 17 | Cite each included study and present its characteristics. | Results: Study characteristics – Page 6; Line 137 - 226 |
| Risk of bias in studies | 18 | Present assessments of risk of bias for each included study. | Results: Risk of bias – Page 27; Line 406 - 426 |
| Results of individual studies | 19 | For all outcomes, present, for each study: (a) summary statistics for each group (where appropriate) and (b) an effect estimate and its precision (e.g. confidence/credible interval), ideally using structured tables or plots. | Results: Predictive performance – Page 19; Line 305 - 358 |
| Results of syntheses | 20a | For each synthesis, briefly summarise the characteristics and risk of bias among contributing studies. | Results: Overall pooled discrimination – Page 19; Line 306 - 329 |
| | 20b | Present results of all statistical syntheses conducted. If meta-analysis was done, present for each the summary estimate and its precision (e.g. confidence/credible interval) and measures of statistical heterogeneity. If comparing groups, describe the direction of the effect. | Results: Overall pooled discrimination – Page 19; Line 306 - 329 |
| | 20c | Present results of all investigations of possible causes of heterogeneity among study results. | Results: Subgroup analyses/Meta-regression – Page 21; Line 330 - 358 |
| | 20d | Present results of all sensitivity analyses conducted to assess the robustness of the synthesized results. | Not reported |
| Reporting biases | 21 | Present assessments of risk of bias due to missing results (arising from reporting biases) for each synthesis assessed. | Results: Publication bias – Page 28; Line 435 - 450 |
| Certainty of evidence | 22 | Present assessments of certainty (or confidence) in the body of evidence for each outcome assessed. | |
| **DISCUSSION** | | | |
| Discussion | 23a | Provide a general interpretation of the results in the context of other evidence. | Discussion – Page 30 to 35 |
| | 23b | Discuss any limitations of the evidence included in the review. | Discussion: Strengths and limitations – Page 34-35 |
| | 23c | Discuss any limitations of the review processes used. | Discussion: Strengths and limitations – Page 34-35 |
| | 23d | Discuss implications of the results for practice, policy, and future research. | Discussion; Conclusion – Page 35 |
| **OTHER INFORMATION** | | | |

### PRISMA 2020 Checklist

| Section and Topic | Item # | Checklist item | Location where item is reported |
|---|---|---|---|
| Registration and protocol | 24a | Provide registration information for the review, including register name and registration number, or state that the review was not registered. | Methods: Study design and registration – Page 36; Lines 615 - 619 |
| | 24b | Indicate where the review protocol can be accessed, or state that a protocol was not prepared. | Methods: Study design and registration – Page 36; Lines 615 - 619 |
| | 24c | Describe and explain any amendments to information provided at registration or in the protocol. | |
| Support | 25 | Describe sources of financial or non-financial support for the review, and the role of the funders or sponsors in the review. | Funding – Page 40; Line 693 - 701 |
| Competing interests | 26 | Declare any competing interests of review authors. | Competing Interests – Page 40; Line 703 |
| Availability of data, code and other materials | 27 | Report which of the following are publicly available and where they can be found: template data collection forms; data extracted from included studies; data used for all analyses; analytic code; any other materials used in the review. | Not included |

*From:* Page MJ, McKenzie JE, Bossuyt PM, Boutron I, Hoffmann TC, Mulrow CD, et al. The PRISMA 2020 statement: an updated guideline for reporting systematic reviews. BMJ 2021;372:n71. doi: 10.1136/bmj.n71.
This work is licensed under CC BY 4.0. To view a copy of this license, visit https://creativecommons.org/licenses/by/4.0/
