## Supplementary material for "AI/ML-based prediction of TB treatment failure: A systematic review and meta-analysis": Table_S1_study_characteristics: Table_S1_study_characteristics.html

|  |  |  |  |  |  |  |  |  |  |  |  |  |
| --- | --- | --- | --- | --- | --- | --- | --- | --- | --- | --- | --- | --- |
| Characteristics of Included Studies | | | | | | | | | | | | |
| All included studies (N = 34) | | | | | | | | | | | | |
| ID | First Author | Year | Country | Study Design | N | Drug\_Resistance | TB\_Site | HIV\_Status | External validation | Best model | Primary AUC | PROBAST |
| 33 | Pusch | 2014 | USA | Retrospective cohort study | 438 | Drug-sensitive | Extrapulmonary | Included | No | CART | NA | High |
| 5 | Modongo | 2016 | Botswana | Prospective clinical study | 28 | Drug-resistant | Pulmonary | Included | No | CART | NA | High |
| 11 | Swaminathan | 2016 | India | Prospective PK/PD study | 161 | Drug-sensitive | Pulmonary and Extrapulmonary | Included | No | Boosted CART | 0.750 | High |
| 7 | Pasipanodya | 2018 | Benin, Guinea, Kenya, Senegal, South Africa | Nested pharmacokinetic (PK) study | 126 | Not specified | Pulmonary | Included | No | Random Forest / Boosted CART | 0.808 | High |
| 16 | Sauer | 2018 | Azerbaijan, Belarus, Georgia, Moldova, Romania | Retrospective Study | 587 | Drug-sensitive | Pulmonary and Extrapulmonary | Not reported | No | Forward stepwise selection | 0.740 | High |
| 24 | Hussain | 2019 | Pakistan | Retrospective Study | 4213 | Drug-sensitive | Pulmonary | Not reported | No | Random Forest | NA | High |
| 14 | Radtke | 2019 | Multinational | Mathematical modeling study | 133302 | Drug-sensitive | Pulmonary and Extrapulmonary | Not reported | No | N/A | NA | High |
| 2 | Asad | 2020 | Azerbaijan, Belarus, Georgia, India, Moldova, Romania | Retrospective / Machine learning framework | 1533 | Drug-resistant | Pulmonary | Not reported | No | Artificial Neural Network (ANN) | NA | High |
| 3 | Singh | 2020 | India | Mathematical transmission dynamics model | 637 | Drug-sensitive | Pulmonary | Not reported | No | N/A | NA | High |
| 26 | Kanesamoorthy | 2021 | Myanmar | Retrospective Study | 356 | Mixed | Pulmonary | Not reported | No | Support Vector Machine (Linear and RBF kernel) | NA | High |
| 30 | Rosenfeld | 2021 | Multinational | Retrospective case-control | 253 | Not specified | Pulmonary | Not reported | No | N/A | NA | High |
| 19 | Kheirandish | 2022 | Moldova | Retrospective Study | 17958 | Drug-sensitive | Pulmonary | Not reported | No | Random Forest | NA | High |
| 28 | Osório | 2022 | Mozambique | Retrospective cohort study | 3012 | Drug-sensitive | Pulmonary and Extrapulmonary | Included | No | Logistic regression | 0.717 | High |
| 18 | Shivakoti | 2022 | India | Case-control study nested within a prospective cohort | 192 | Drug-sensitive | Pulmonary | Included | No | Random Forest | 0.790 | High |
| 12 | Zheng | 2022 | China | Multicentre prospective cohort study | 197 | Drug-resistant | Pulmonary | Excluded | No | CART (Classification and Regression Tree) | NA | High |
| 29 | Anley | 2023 | Ethiopia | Multicenter retrospective follow-up study | 517 | Drug-resistant | Pulmonary and Extrapulmonary | Included | No | Multivariable Logistic regression risk score | 0.793 | High |
| 8 | Nijiati | 2023 | China | Retrospective Study | 284 | Not specified | Pulmonary | Not reported | Yes | Fusion of Bagging and SDLM | 0.802 | High |
| 9 | Nijiati | 2023 | China | Retrospective Study | 579 | Not specified | Pulmonary | Not reported | Yes | PM3 (Pretreatment + 2 follow-up scans) | 0.815 | High |
| 21 | Fayaz | 2024 | India | Retrospective clinical trial study | 1236 | Drug-sensitive | Pulmonary | Not reported | No | Decision Tree | 0.909 | High |
| 25 | Hosu | 2024 | South Africa | Retrospective review | 456 | Drug-resistant | Pulmonary and Extrapulmonary | Included | No | Decision tree classifier | NA | High |
| 6 | Kim | 2024 | South Korea | Retrospective multicenter cohort study | 230 | Not specified | Pulmonary | Not reported | No | Multivariate logistic regression | NA | High |
| 23 | Li | 2024 | Multinational | Retrospective multi-cohort analysis | 1728 | Mixed | Pulmonary | Not reported | Yes | Neural Network | NA | High |
| 15 | Peng | 2024 | China | Retrospective Study | 429 | Drug-sensitive | Pulmonary | Not reported | No | XGBoost | 0.928 | High |
| 22 | Rodrigues | 2024 | Brazil | Retrospective cohort study | 243726 | Drug-sensitive | Pulmonary | Included | No | Light Gradient Boosting | 0.720 | Low |
| 20 | Sambarey | 2024 | Multinational | Retrospective multimodal analysis | 4139 | Drug-sensitive | Pulmonary | Not reported | Yes | Multimodal Random Forest | 0.840 | High |
| 4 | Yang | 2024 | China | Multicenter Retrospective Study | 524 | Drug-resistant | Pulmonary | Not reported | No | Logistic Regression Nomogram | 0.829 | High |
| 1 | Öhrnberg | 2024 | Peru, Kenya | Longitudinal pilot and validation study | 71 | Drug-sensitive | Pulmonary | Included | Yes | Elastic net regression | NA | High |
| 10 | Lu | 2025 | China | Retrospective inpatient study | 1625 | Mixed | Pulmonary | Excluded | No | Integrated Machine Learning Model / Logistic regression nomogram | 0.815 | High |
| 17 | Lv | 2025 | China | Retrospective multicenter study | 297 | Drug-resistant | Pulmonary | Excluded | Yes | Transformer Fusion Model | 0.879 | High |
| 31 | Phat | 2025 | Brazil | Retrospective cohort study | 665883 | Mixed | Pulmonary and Extrapulmonary | Not reported | No | Random Forest | NA | High |
| 13 | Rojas-Carabali | 2025 | Multinational | Retrospective longitudinal study | 2023 | Mixed | Extrapulmonary | Not reported | No | XGBoost (6-month) / Random Forest (12-month) | 0.915 | High |
| 27 | Wang | 2025 | China | Retrospective cohort study | 1086 | Mixed | Pulmonary | Excluded | No | K-Nearest Neighbors (KNN) | 0.870 | High |
| 32 | Zhang | 2025 | China | Retrospective cohort study | 240 | Drug-resistant | Pulmonary | Excluded | Yes | Random Forest | 0.998 | High |
| 34 | Zhang | 2025 | China | Retrospective cohort study | 881 | Drug-resistant | Pulmonary | Not reported | Yes | Artificial Neural Network (ANN) | 0.900 | High |
