## Supplementary material for "AI/ML-based prediction of TB treatment failure: A systematic review and meta-analysis": Table_S2_probast_study_level: Table_S2_probast_study_level.html

|  |  |  |  |  |  |  |  |
| --- | --- | --- | --- | --- | --- | --- | --- |
| **Study-level PROBAST assessment and reasons for risk of bias** | | | | | | | |
| Detailed study-level judgments across PROBAST domains | | | | | | | |
| Study ID | Study | Participants | Predictors | Outcome | Analysis | Overall risk | Key reason(s) for bias |
| 1 | Öhrnberg 2024 | Low | Low | Low | High | High | Analysis: Lack of calibration assessment reported. |
| 2 | Asad 2020 | Low | Low | Low | High | High | Analysis: Lack of calibration assessment reported.; Analysis: Feature selection based on univariable analysis (e.g., p-value screening). |
| 3 | Singh 2020 | Low | Low | Low | High | High | Analysis: Lack of adequate internal or external validation. |
| 4 | Yang 2024 | Low | Low | Low | High | High | Analysis: Inappropriate missing data handling (complete case analysis/exclusion). |
| 5 | Modongo 2016 | Low | Low | Low | High | High | Analysis: Lack of calibration assessment reported. |
| 6 | Kim 2024 | Low | Low | Low | High | High | Analysis: Inappropriate missing data handling (complete case analysis/exclusion).; Analysis: Lack of calibration assessment reported.; Analysis: Lack of adequate internal or external validation.; Analysis: Feature selection based on univariable analysis (e.g., p-value screening). |
| 7 | Pasipanodya 2018 | Low | Low | Low | High | High | Analysis: Inappropriate missing data handling (complete case analysis/exclusion).; Analysis: Lack of calibration assessment reported. |
| 8 | Nijiati 2023 | Low | Low | Low | High | High | Analysis: Lack of calibration assessment reported. |
| 9 | Nijiati 2023 | Low | Low | Low | High | High | Analysis: Lack of calibration assessment reported. |
| 10 | Lu 2025 | Low | Low | Low | High | High | Analysis: Inappropriate missing data handling (complete case analysis/exclusion).; Analysis: Feature selection based on univariable analysis (e.g., p-value screening). |
| 11 | Swaminathan 2016 | Low | Low | Low | High | High | Analysis: Inappropriate missing data handling (complete case analysis/exclusion).; Analysis: Lack of calibration assessment reported. |
| 12 | Zheng 2022 | Low | Low | Low | High | High | Analysis: Lack of calibration assessment reported.; Analysis: Lack of adequate internal or external validation. |
| 13 | Rojas-Carabali 2025 | Low | Low | Low | High | High | Analysis: Lack of calibration assessment reported. |
| 14 | Radtke 2019 | Low | Low | Low | High | High | Analysis: Lack of calibration assessment reported.; Analysis: Lack of adequate internal or external validation. |
| 15 | Peng 2024 | Low | Low | Low | High | High | Analysis: Inappropriate missing data handling (complete case analysis/exclusion).; Analysis: Lack of calibration assessment reported. |
| 16 | Sauer 2018 | Low | Low | Low | High | High | Analysis: Lack of calibration assessment reported. |
| 17 | Lv 2025 | Low | Low | Low | High | High | Analysis: Inappropriate missing data handling (complete case analysis/exclusion).; Analysis: Lack of calibration assessment reported. |
| 18 | Shivakoti 2022 | High | Low | Low | High | High | Participants: Case-control design introduces potential selection bias.; Analysis: Lack of calibration assessment reported. |
| 19 | Kheirandish 2022 | Low | Low | Low | High | High | Analysis: Lack of calibration assessment reported. |
| 20 | Sambarey 2024 | Low | Low | Low | High | High | Analysis: Lack of calibration assessment reported. |
| 21 | Fayaz 2024 | Low | Low | Low | High | High | Analysis: Lack of calibration assessment reported. |
| 22 | Rodrigues 2024 | Low | Low | Low | Low | Low | Not specified |
| 23 | Li 2024 | Low | Low | Low | High | High | Analysis: Lack of calibration assessment reported. |
| 24 | Hussain 2019 | Low | Low | Low | High | High | Analysis: Lack of calibration assessment reported. |
| 25 | Hosu 2024 | Low | Low | Low | High | High | Analysis: Lack of calibration assessment reported. |
| 26 | Kanesamoorthy 2021 | Low | Low | Low | High | High | Analysis: Lack of calibration assessment reported. |
| 27 | Wang 2025 | Low | Low | Low | High | High | Analysis: Lack of calibration assessment reported. |
| 28 | Osório 2022 | Low | Low | Low | High | High | Analysis: Inappropriate missing data handling (complete case analysis/exclusion).; Analysis: Lack of adequate internal or external validation.; Analysis: Feature selection based on univariable analysis (e.g., p-value screening). |
| 29 | Anley 2023 | Low | Low | Low | High | High | Analysis: Feature selection based on univariable analysis (e.g., p-value screening). |
| 30 | Rosenfeld 2021 | High | Low | Low | High | High | Participants: Case-control design introduces potential selection bias.; Analysis: Lack of calibration assessment reported. |
| 31 | Phat 2025 | Low | Low | Low | High | High | Analysis: Inappropriate missing data handling (complete case analysis/exclusion).; Analysis: Lack of calibration assessment reported. |
| 32 | Zhang 2025 | Low | Low | Low | High | High | Analysis: Lack of calibration assessment reported. |
| 33 | Pusch 2014 | Low | Low | Low | High | High | Analysis: Lack of calibration assessment reported. |
| 34 | Zhang 2025 | Low | Low | Low | High | High | Analysis: Inappropriate missing data handling (complete case analysis/exclusion).; Analysis: Lack of calibration assessment reported.; Analysis: Feature selection based on univariable analysis (e.g., p-value screening). |
|  |  |  |  |  |  |  |  |
| --- | --- | --- | --- | --- | --- | --- | --- |
| **Abbreviation:** PROBAST, Prediction model Risk Of Bias Assessment Tool. | | | | | | | |
| **Note:** Overall risk of bias reflects the highest risk judgment across domains. This table provides the detailed study-level assessments underlying the summary shown in Table 10 and Figure 13. | | | | | | | |
