## Supplementary material for "AI/ML-based prediction of TB treatment failure: A systematic review and meta-analysis": Search strategy

(

    "Tuberculosis, Pulmonary"[Mesh] OR "Mycobacterium tuberculosis"[Mesh] OR

    "Pulmonary Tuberculosis"[Title/Abstract] OR "Tuberculosis"[Title/Abstract] OR

    "TB"[Title/Abstract] OR "Active TB"[Title/Abstract] OR "PTB"[Title/Abstract]

)

AND

(

    "Artificial Intelligence"[Mesh] OR "Machine Learning"[Mesh] OR "Deep Learning"[Mesh] OR

    "Neural Networks, Computer"[Mesh] OR "Artificial Intelligence"[Title/Abstract] OR

    "Machine Learning"[Title/Abstract] OR "Deep Learning"[Title/Abstract] OR

    "Neural Network*"[Title/Abstract] OR "Random Forest"[Title/Abstract] OR

    "Support Vector Machine"[Title/Abstract] OR "SVM"[Title/Abstract] OR

    "Decision Tree"[Title/Abstract] OR "Gradient Boosting"[Title/Abstract] OR

    "XGBoost"[Title/Abstract] OR "CNN"[Title/Abstract] OR "Computer Vision"[Title/Abstract]

)

AND

(

    "Treatment Outcome"[Mesh] OR "Prognosis"[Mesh] OR "Treatment Failure"[Title/Abstract] OR

    "Treatment Success"[Title/Abstract] OR "Sputum conversion"[Title/Abstract] OR

    "Non-conversion"[Title/Abstract] OR "Relapse"[Title/Abstract] OR

    "Recurrence"[Title/Abstract] OR "Mortality"[Title/Abstract] OR

    "Drug Resistance"[Title/Abstract] OR "MDR-TB"[Title/Abstract] OR

    "Predict*"[Title/Abstract] OR "Prognos*"[Title/Abstract]

)
